# Archetypal analysis of COVID-19 in Montana, USA, March 13, 2020 to April 26, 2022

**DOI:** 10.1101/2023.03.06.23286886

**Authors:** Emily Stone, Sebastian Coombs, Erin Landguth

## Abstract

Given the potential consequences of infectious diseases, it is important to understand how broad scale incidence variability influences the probability of localized outbreaks. Often, these infectious disease data can involve complex spatial patterns intermixed with temporal trends. Archetypal Analysis is a method to mine complex spatiotemporal epidemiological data, and can be used to discover the dynamics of spatial patterns. The application of Archetypal Analysis to epistemological data is relatively new, and here we present one of the first applications using COVID-19 data from March 13, 2020 to April 26, 2022, in the counties of Montana, USA. We present three views of the data set with Archetypal Analysis. First, we evaluate the entire 56 county data set. Second, we compute mutual information of the 56 counties’ time series to remove counties whose dynamics are mainly independent from most of the other counties. We choose the top 17 counties ranked in terms of increasing total mutual information. Finally, to compare how population size might influence results, we conducted an analysis with 10 of the largest counties. Using the Archetypal Analysis results, we analyze the disease outbreaks across Montana, comparing and contrasting the three different cases and showing how certain counties can be found in distinct sets of archetypes. Using the reconstruction time series, we show how each outbreak had a unique trajectory across the state in terms of the archetypes.

**Author summary:** Archetypal Analysis provides an additional tool for the study of spatio-temporal epidemiological data. We apply Archetypal Analysis to COVID-19 data and reveal how this approach can be used to analyse the dynamics of each COVID-19 outbreak across the state.

## Introduction

The COVID-19 pandemic launched an intensive effort to understand the processes and drivers of infectious disease outbreaks, with a growing emphasis on improving predictions and providing information to help mitigate public health threats. [12] The unprecedented collection of spatially dense data from across the world, along with increased computing power and new implementations, has made spatio-temporal approaches tractable. Combining spatial with temporal methods allows an investigator to not only study the persistence of patterns over time, but study the evolution of these pattern. The patterns themselves may be unexpected and the methods can detect spatial clustering could may reveal environmental causatives or potential data recording errors.

Archetypal Analysis (AA) is a promising data mining tool for epidemiological data and the analyses of the spatio-temporal spread of disease. AA attempts to capture patterns within such multivariate data sets, and use these to represent the time evolution of the spatial data. The archetypal patterns are convex combinations of the data points themselves, and as such resemble the data, making interpretation much more transparent than linear decompositions like principal component analysis (PCA) [1]. Cutler and Breiman introduced AA as variant of PCA that could capture ‘archetypal patterns’ in the data [7]. More specifically, each time-based observation can be constructed as a convex combination of the archetypes, representing the set with a limited number of points on the convex hull. PCA provides the best way to compress data, but the eigenvectors produced are not and should not be interpreted as actual data points, they are directions in the high dimensional space being decomposed. The archetypes, along with the reconstruction time series, represent the data as moving from one representative observation to another as the epidemic evolves. In clustered data, AA typically finds the centroids of clusters, and represents data as convex combinations of the clusters, so it can indicate if a point lies between several clusters. Therefore, AA combines the strengths of other commonly used techniques for data decomposition; providing the interpretability that PCA lacks and more flexibility than many clustering algorithms (i.e. it is a soft clustering algorithm).

AA applications have spanned many fields, and include the analysis of weather and climate patterns [6, 11, 22, 24], machine learning [16], market analysis [13], and biomedical and industrial engineering [9, 26]. Mokhtari et al. [4] use AA in one of the first applications to epidemiological data, reconstructing the spatio-temporal patterns in an influenza time series, and showing how prominent outbreaks developed across space and for each influenza season (2010-2019). Here, we follow their approach and apply AA to COVID-19 county-level data in Montana. Our goal is to further evaluate the use of AA and attempt to reconstruct the spatial patterns for each COVID-19 outbreak from March 2020 to April 2022. The full 56 dimension data set is decomposed by AA, along with 2 reduced dimension sets, one reduced using a mutual information metric (17-dimensional), and another considering only counties with large population centers (10-dimensional). We apply AA to decompose them into a limited number of spatial patterns of disease counts in each county for the specific outbreaks. The patterns found in each outbreak can be compared, and a reconstruction coefficient time series allows examination of the spread of COVID-19 from one pattern to another.

We would like to note here a common misconception in the later papers applying Archetypal Analysis. The Archetype algorithm finds points that are convex combinations of the data points that minimize the error in a reconstruction of the data in terms of convex combinations of the archetypes. Recent publications use the term “extremes” [6] to mean archetypes, which is indeed the case if there are significant outliers in the data set. Archetypal decomposition is sensitive to outliers, so “extremes” may be found first. However, this depends strongly on how the data are distributed, especially in high dimensional spaces. The error is the residual sum of squares, so if a point occurs often in a data set, the error in reproducing it will be multiplied by the number of occurrences of that point, and the algorithm will use it as an archetype, necessarily. It may or may not be an “extreme” in the data set. Calling Archetypes “extremes” is a muddy issue, and we discuss this in detail for our data set in what follows.

## Methods

### COVID-19 data for Montana, USA

For our analysis, we are presented with COVID-19 data from counties (*m* = 56; i.e., spatial attributes) in Montana, USA, courtesy of Montana Department of Health and Human Services from March 13, 2020 - April 26, 2022. This covers 110 full weeks or *n* = 776 daily time observations. The first case was observed on March 13, 2020, however, we removed the first 100 days because Montana was following non-pharmaceutical interventions during this time and did not experience the pandemic’s initial wave observed in the rest of the country. Therefore, we analyzed the period from June 20, 2020 – April 26, 2022, resulting in *n* = 676 observations. During this time period, Montana experienced three peaks in cases followed by declines (i.e., outbreaks or waves). A running weekly average of COVID-19 cases per day were smoothed using the “smoothdata” function within MATLAB (default setting of mean = 3) to reduce noise in the time series. Fig. 1 shows the data reported for each county. To account for differences in population size, we weighted the COVID-19 cases for each county by the population size of the county in 2020 (American Community survey data from the Census Bureau) and reported cases per 1000 people in that county.

**Fig 1.**
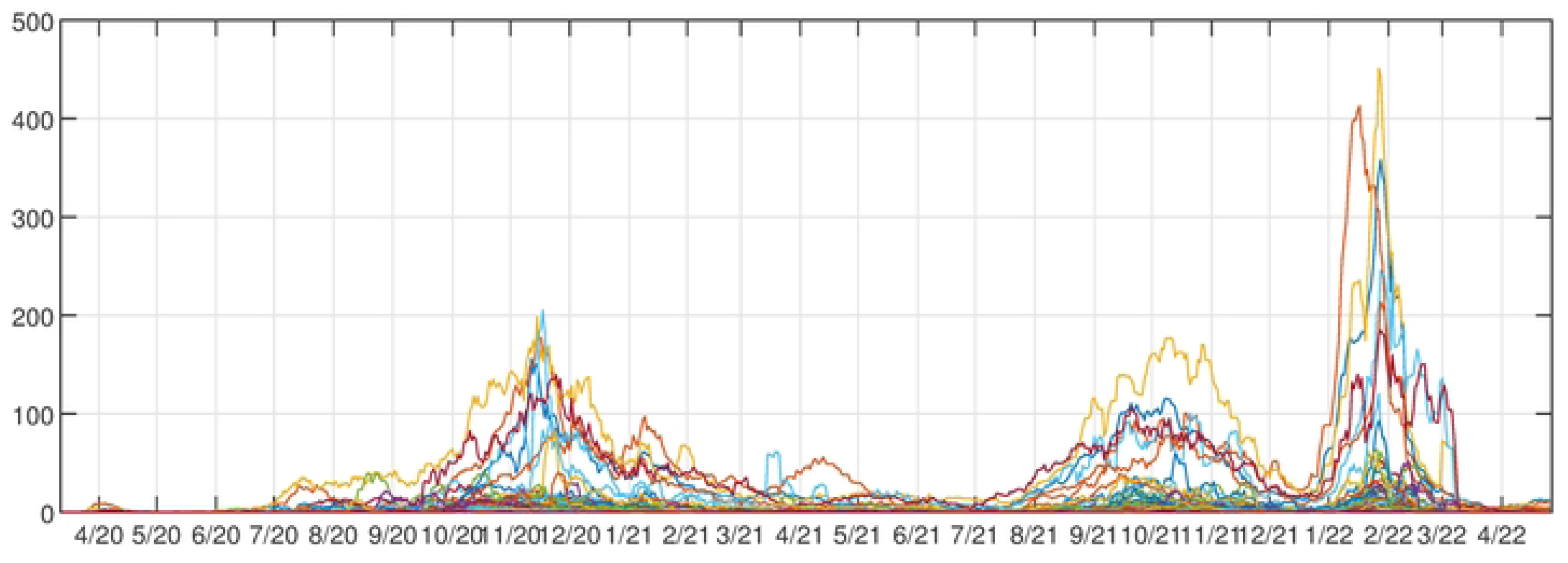
Montana County Cases. Running weekly average of COVID-19 cases plotted for all Montana counties.

**Fig 2.**
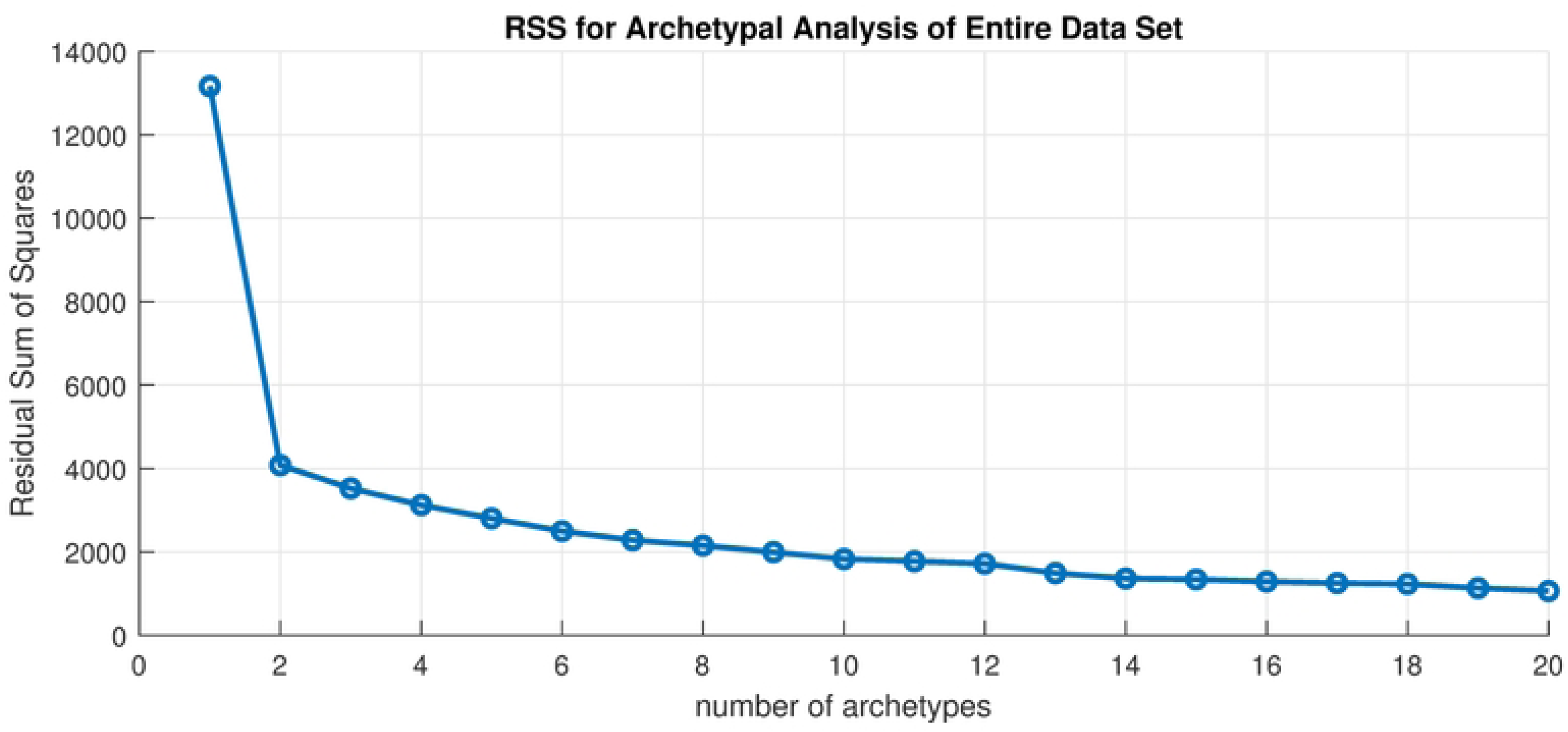
Scree plot. Residual sum of squares vs. number of archetypes in the set.

### Archetypal Analysis: Mathematical Formulation

Consider an *m × n* matrix **X**, where *n* is the number of observations of COVID-19 cases across *m* Montana counties. AA decomposes the spatio-temporal variability of **X** in a similar way to PCA but with the following underlying constraints. Given a specified value for *k*, AA identifies *m*-dimensional vectors **z**_1_*,‧ ‧ ‧* **z_k_** that best describe *k* characteristic patterns, or archetypes, in the original data set, such that data can be represented as convex combinations (i.e., linear combinations with non-negative coefficients that sums to unity) of these archetypal patterns:

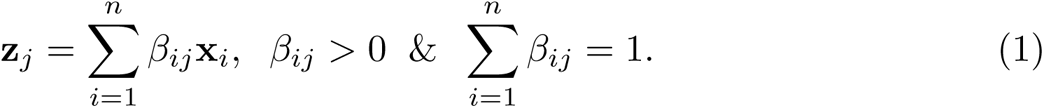

The *n*-dimensional vector *β_j_* contains the convex weights for the *j*th archetype across all observations. The *n × k* matrix of all such weights is given by **B** = *β*_1_ *‧‧‧ β_k_*. Each archetype is either a convex combinations of the original observations or an actual observation [7], so they are more readily interpreted compared to PCA eigenvectors. All observations can then be approximated by a convex combination of the archetypes:

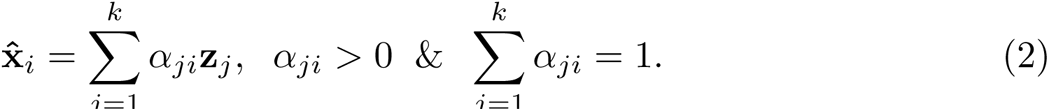

Here, the convex weights, sometimes referred to as mixture coefficients, *α_ji_* with *j* = 1, · · · *, k* range from 0 to 1, are used to reconstruct the *i*th observation across the *k* archetypes. The *k* × *n* matrix of all such weights is given by **A** = {*α*_1_, · · · *, α_n_*}. The *α_j_* are like the (nonlinear) projection of the original data **X** onto the *j*th archetype **z***_j_*, similar to PC scores in PCA. Thus the *α_j_*s are time series that determine how much of each archetype is used in reconstructing each data point.

The *m* × *k* matrix **Z** of *k* archetypes is defined by the matrix factorization problem:

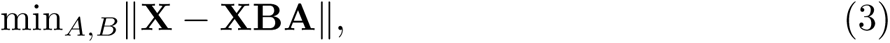

where **Z** = **XB**. *RSS* = ||**X** − **XBA**|| is the residual sum of square errors, where ||.|| is the spectral norm. AA seeks to find *k m*-dimensional archetypes such that the *RSS* is minimized. This approach is described in detail in [7], but can be summarized as follows: AA uses a convex least-squares method (CLSM) to estimate the coefficient *α_ji_*, subject to the constraints for given some initial values of *β_ij_*. It then finds the best *β_ij_* using CLSM, using the new *α_ji_*. This process repeats until the RSS fails to improve, or potentially until the maximum number of iterations is reached. AA will find local minimums, not necessarily the global minimum of RSS, hence using several starting *β_ij_* values to insure a global solution is recommended. Furthermore, there is no universal method for determining the optimal value of *k*. One commonly used approach is the “elbow” criteria, where a good value of *k* is selected by when the RSS fails to improve, which can be determined by finding an elbow in the relationship between RSS and *k* in a scree plot. Since its introduction, other algorithms have been developed to find an archetypal decomposition of data. To compute the archetypes we used Matlab packages by Morten Mørup and Lars Kai Hansen [16] for computing the Principal Convex Hull [2]. Archetypes themselves are presented as color mapped counties within a state map, and are created with GeoPandas (GeoPandas.org).

It is noted in [7] that the archetypal points **z**, viewed as vectors, are not orthogonal and have no natural nesting structure, i.e., as more archetypes are found, the archetypes in the smaller set can change. This is in contrast to PCA, where the set of the leading *N* principal components are a subset of the set of the leading *M* principal components for *M > N*. In PCA all the eigenvectors are found in a single decomposition, and computing it is fast and efficient. This is the result of the linearity of PCA, and it comes at the cost of interpretation.

### Mutual Information

To reduce the dimension of the data set, we apply information-theoretic measures introduced by Shannon [21] to quantify the dependence of the count time series from different counties upon each other. For instance, if a county has a COVID-19 count time series that runs more or less independently of the other counties (as was typical with very small population counties), we could chose to remove them from the data set, thereby reducing the dimension. The exact meaning of “more or less independently” is explored below.

Mutual information measures the expected reduction in uncertainty about *x* that results from learning *y*, or vice versa, where *x* and *y* are samples of the random variables *X* and *Y*. This quantity can be formulated

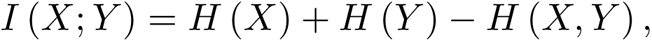

where **entropy** is defined

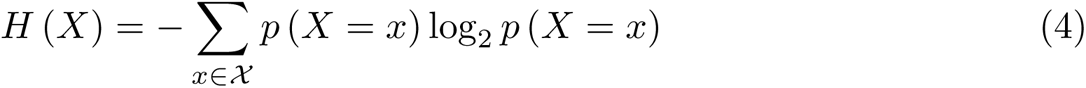

and the **joint entropy** of two random variables *X* and *Y* quantifies the uncertainty of their joint distribution.

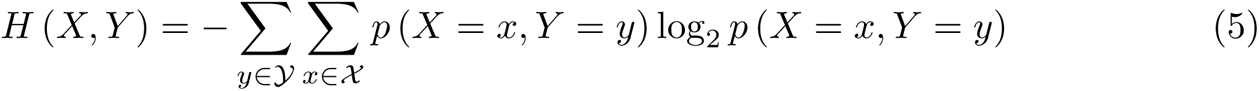

Using Eqs. (4) and (5), the mutual information can be rewritten

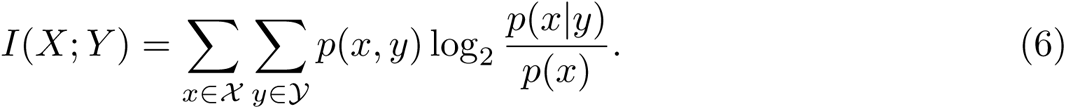

The mutual information is symmetric in the variables *X* and *Y*, *I*(*X*; *Y*) = *I*(*Y*; *X*), and is zero if the random variables are independent or if the relation between them is deterministic (nothing to be learned in either case). Note also that if *X* is statistically correlated to *Y*, *H*(*X|Y*) will be less than *H*(*X*), and *I* will be greater than 0. If *X* is independent of *Y*, *H*(*X|Y*) = *H*(*X*) and *I* = 0. If *X* is uniquely determined by *Y*, *H*(*X|Y*) = 0 and *I*(*X*; *Y*) = *H*(*X*).

In general, association measures like correlation coefficient or mutual information are used to estimate the relationships between two random variables. Correlation coefficient measures such as Pearson or Spearman entail the assumption of linear dependence. Therefore, if two random variables are associated by a nonlinear relationship these methods fail to detect this link, or the strength will be wrongly estimated. Mutual information, however, is able to detect both linear and nonlinear dependencies, and it measures the amount of information connecting two random variables, in this case between disease cases in two Montana counties. In other words, it estimates the reduction in uncertainty about the COVID-19 activity of one county when the activity of another county is known.

## Results

### Archetypal Analysis of 56 Dimension Data Set

As mentioned in an earlier section, we normalized the time series by dividing by the population of each county in 2020 and multiplying it by 1000. This seems like a natural step, and indeed it is customary practice, but in a sparsely populated state like Montana (14 of 56 counties have population less than 5000, and 4 have populations less than 1000) it can lead to issues with our decomposition, as we shall demonstrate now.

We first apply archetypes to the entire truncated in time data set, which is 676 (*m*) observations in 56 (n) dimensions. To determine how many archetypes to compute we consider the scree plot, or RSS versus number of archetypes, plotted in 2. To choose the set of the archetype set, the “elbow criterion” would indicate truncating to one archetype, but this is the “no-disease” archetype that appears in all the decompositions, and serves to turn the counts off and on. Instead, we choose k = 10 archetypes. We are left with a 56 by 10 (n x k) set of *β* values, and a 10 by 676 (k x m) set of *α* values. These 10 archetypes are shown in 3 as color-mapped counties in Montana. The color is determined by the *β* value for that county, for that archetype. Later we refer to high count/large outbreaks as those above 1.5, mid-size outbreaks as between 0.75-1.5, and low level outbreaks as *β* values below 0.75. Each archetypal map can be thought of as a representation of the spatial features of the outbreak at a given time, or period of time. The information in each archetype are summarized in Table 3.

This 10 archetype set is dominated by those with large outbreaks in small population counties. Only two (numbers 8 and 10, Fig. 3 h and j) would be used to represent large outbreaks in the largest population counties (Yellowstone, Missoula, Gallatin, Flathead, Cascade, Lewis and Clark, Ravalli, Silver Bow, Lake and Lincoln). Here the problem with normalizing disease counts by population becomes apparent. Small population counties will have much larger normalized counts on some occasions than all the rest, and these data points are thus outliers. Such small population counties are numerous, and those with population below 10000, in decreasing order from 9391 to 434, are: Beaverhead (at 9391), Deer Lodge, Dawson, Stillwater, Madison, Rosebud, Valley, Blaine, Powell, Broadwater, Teton, Pondera, Chouteau, Tolle, Musselshell, Minderal Phillips, Sweet Grass, Sheridan, Granite, Fallon, Wheatland, Liberty, Judith Basin, Meagher, McCone, Powder River, Daniels, Carter, Prairie, Wibaux, Garfield, Golden Valley, Treasure, Pertoleum (at 434).

**Fig 3.**
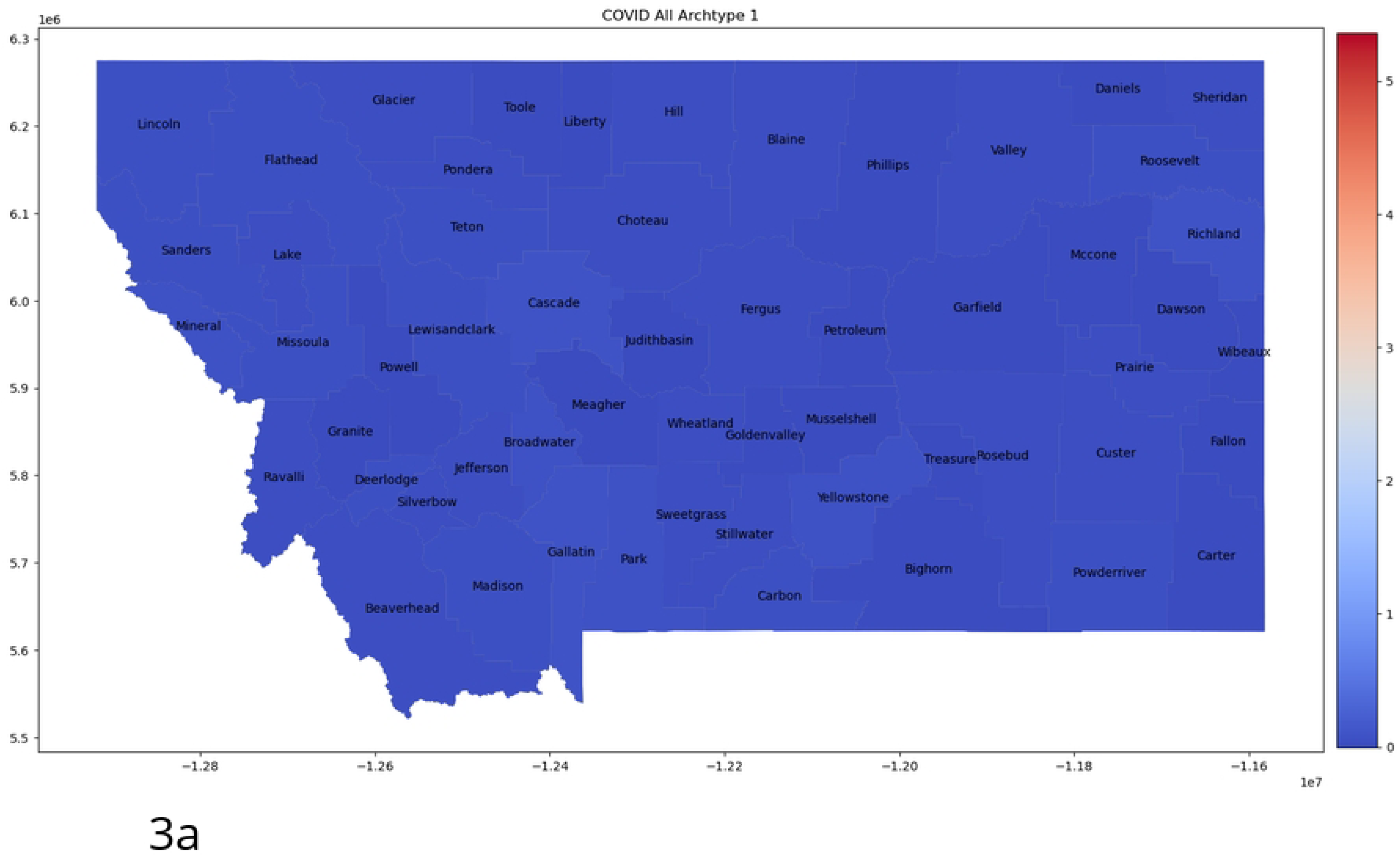
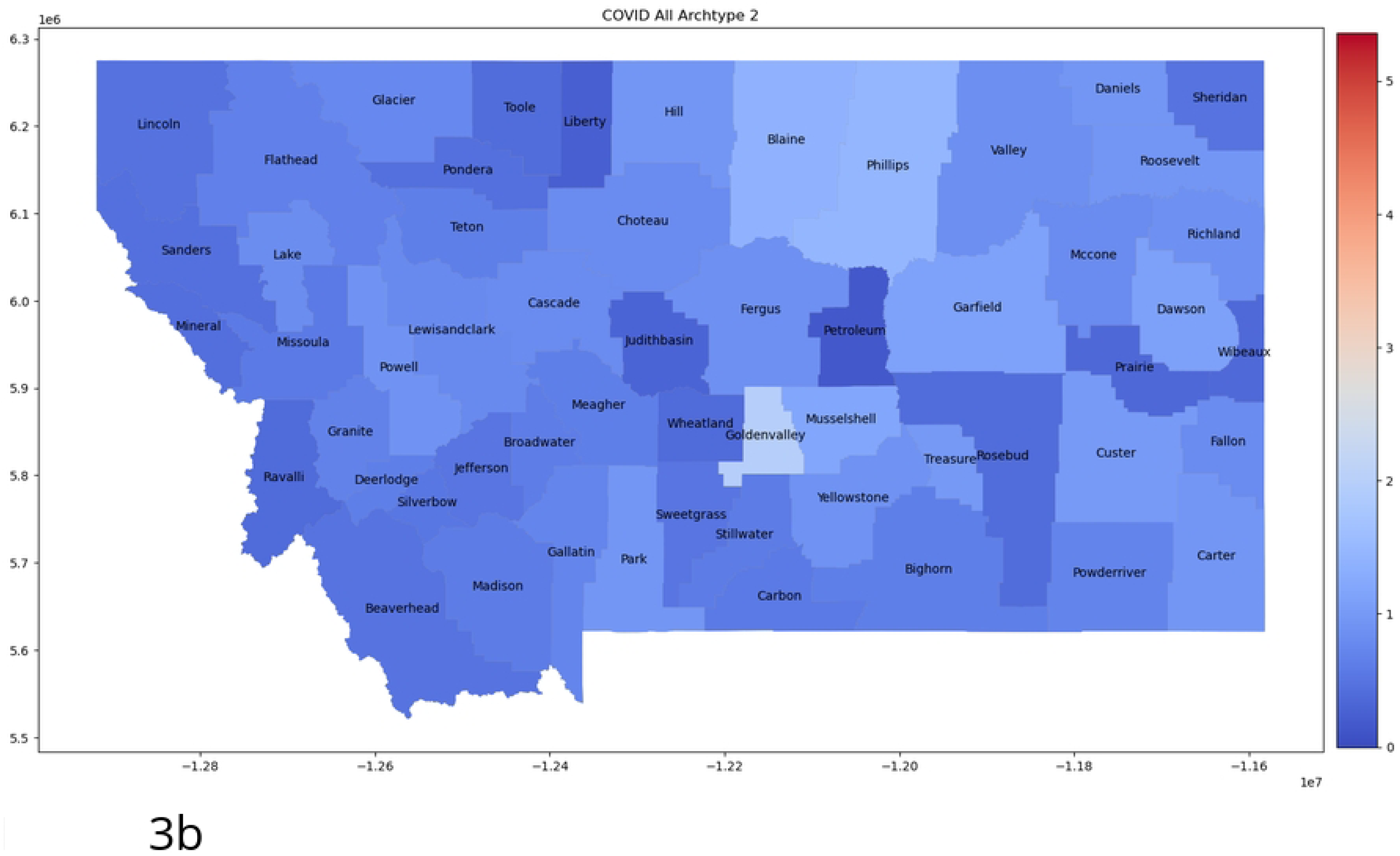
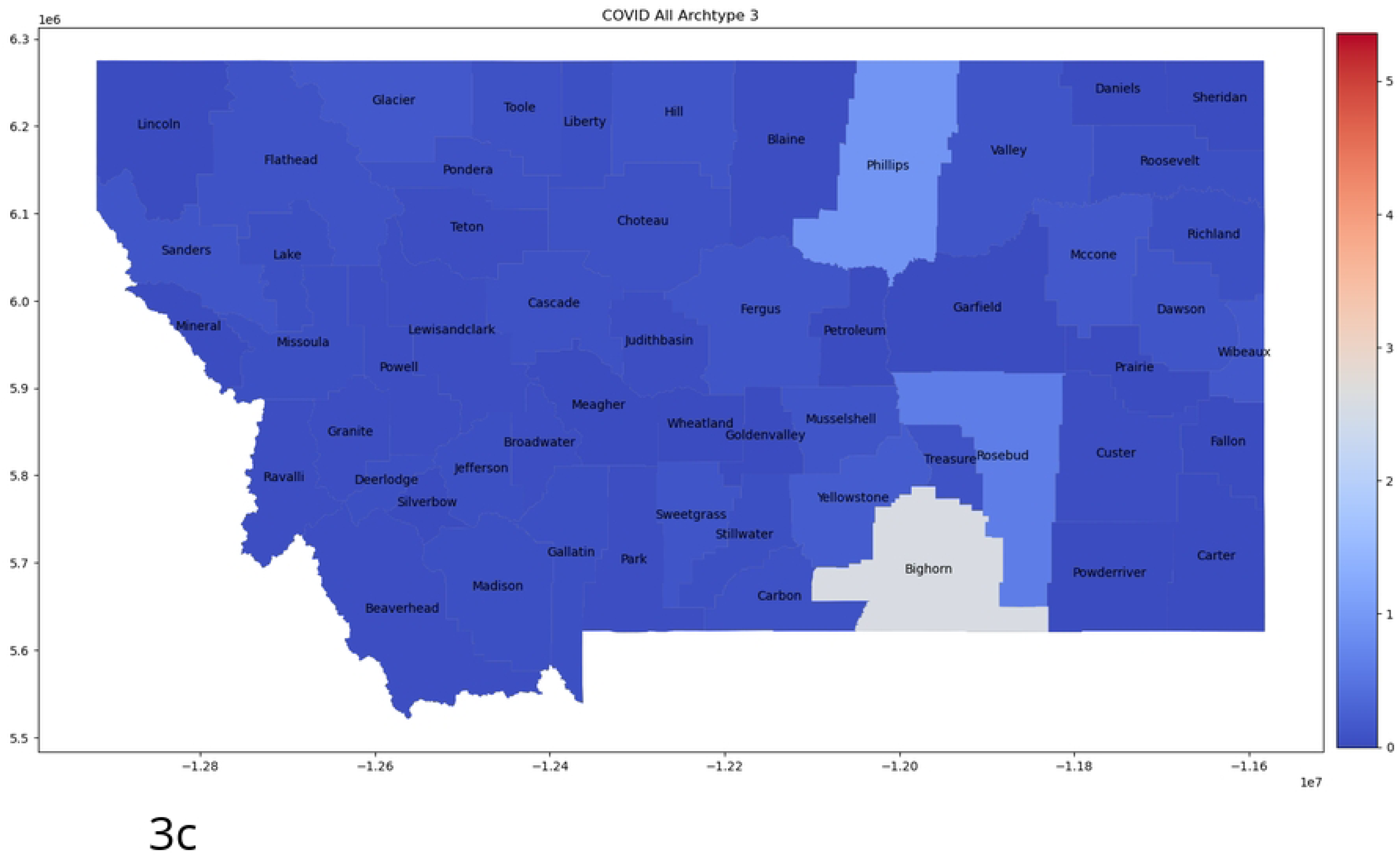
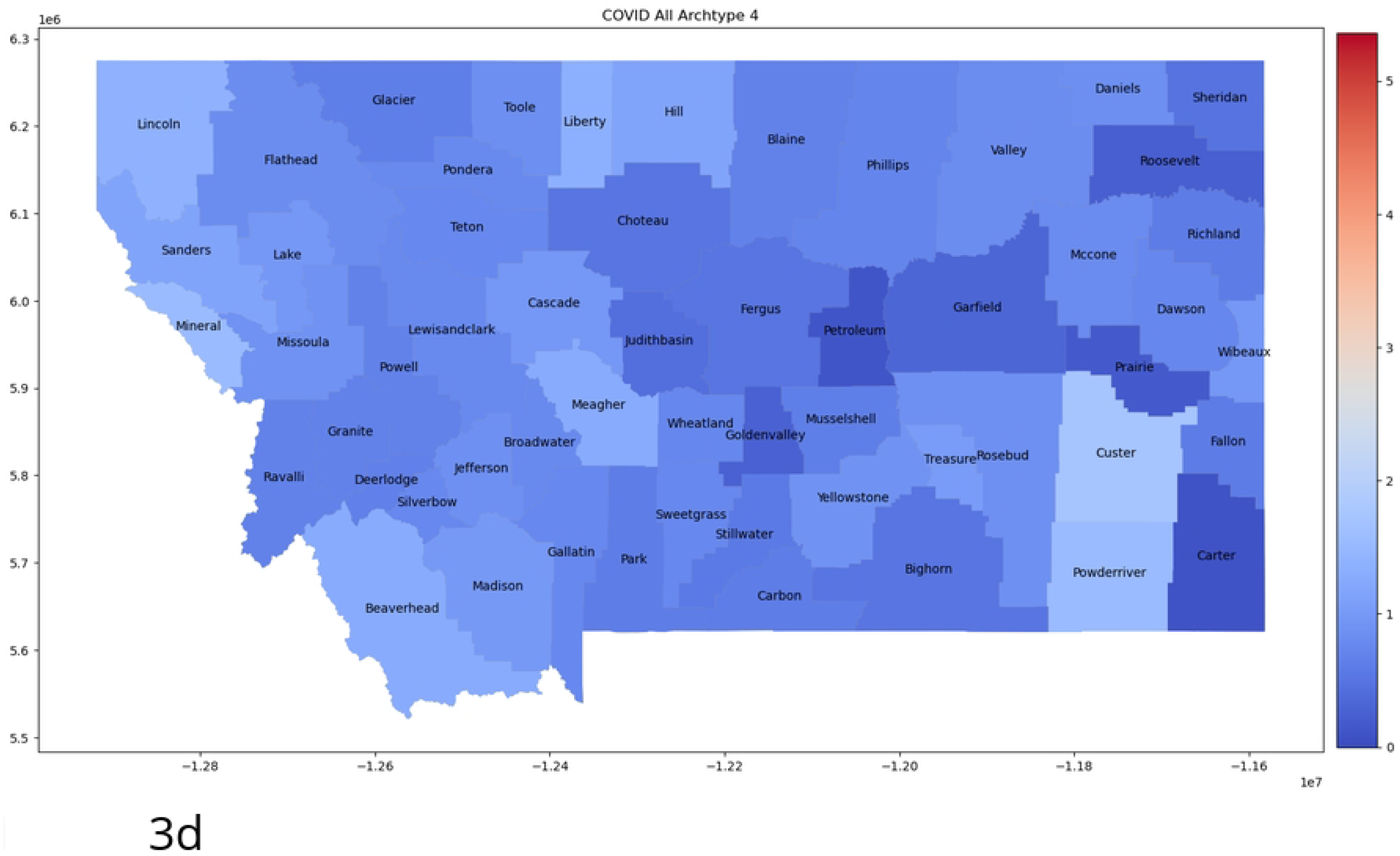
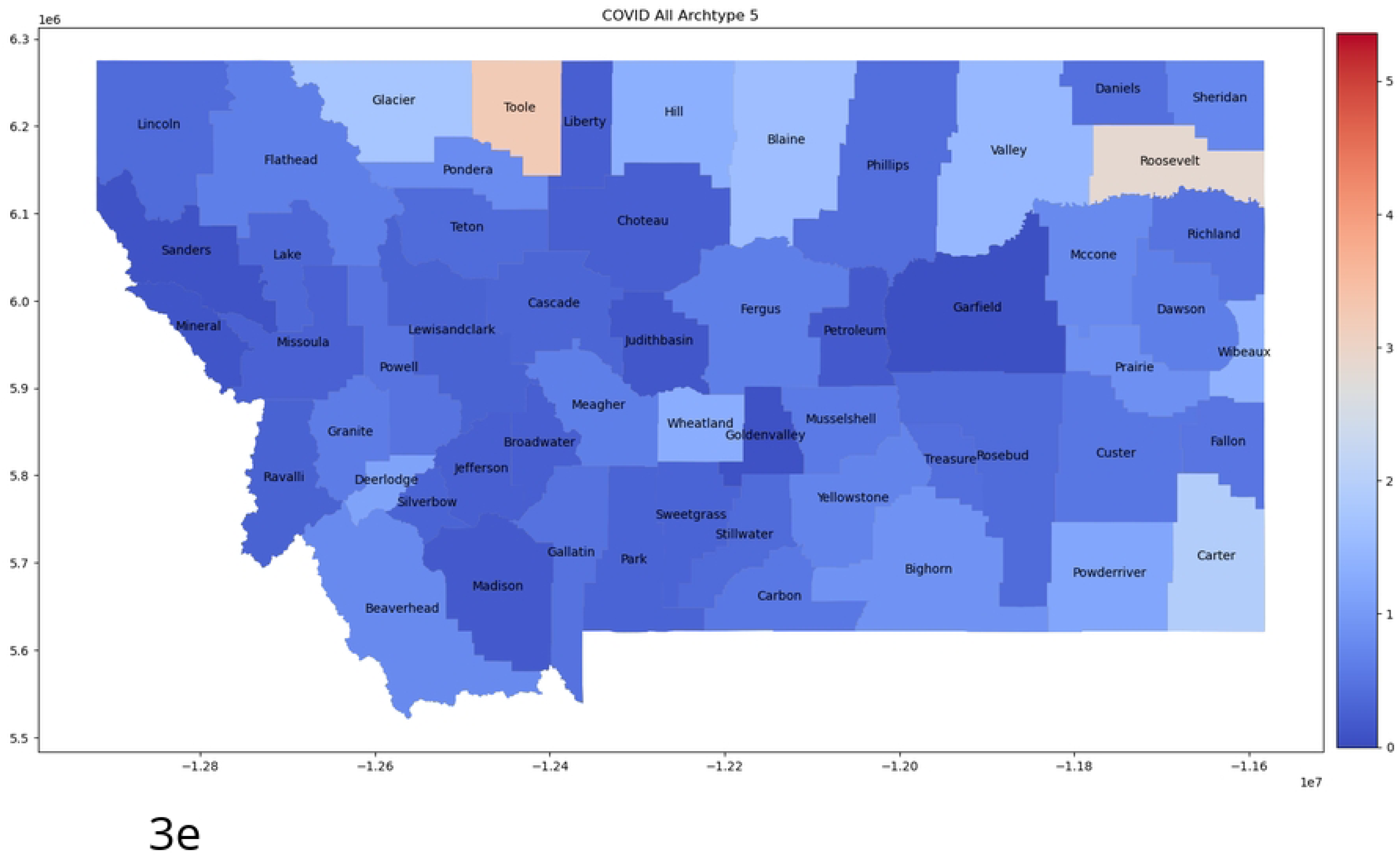
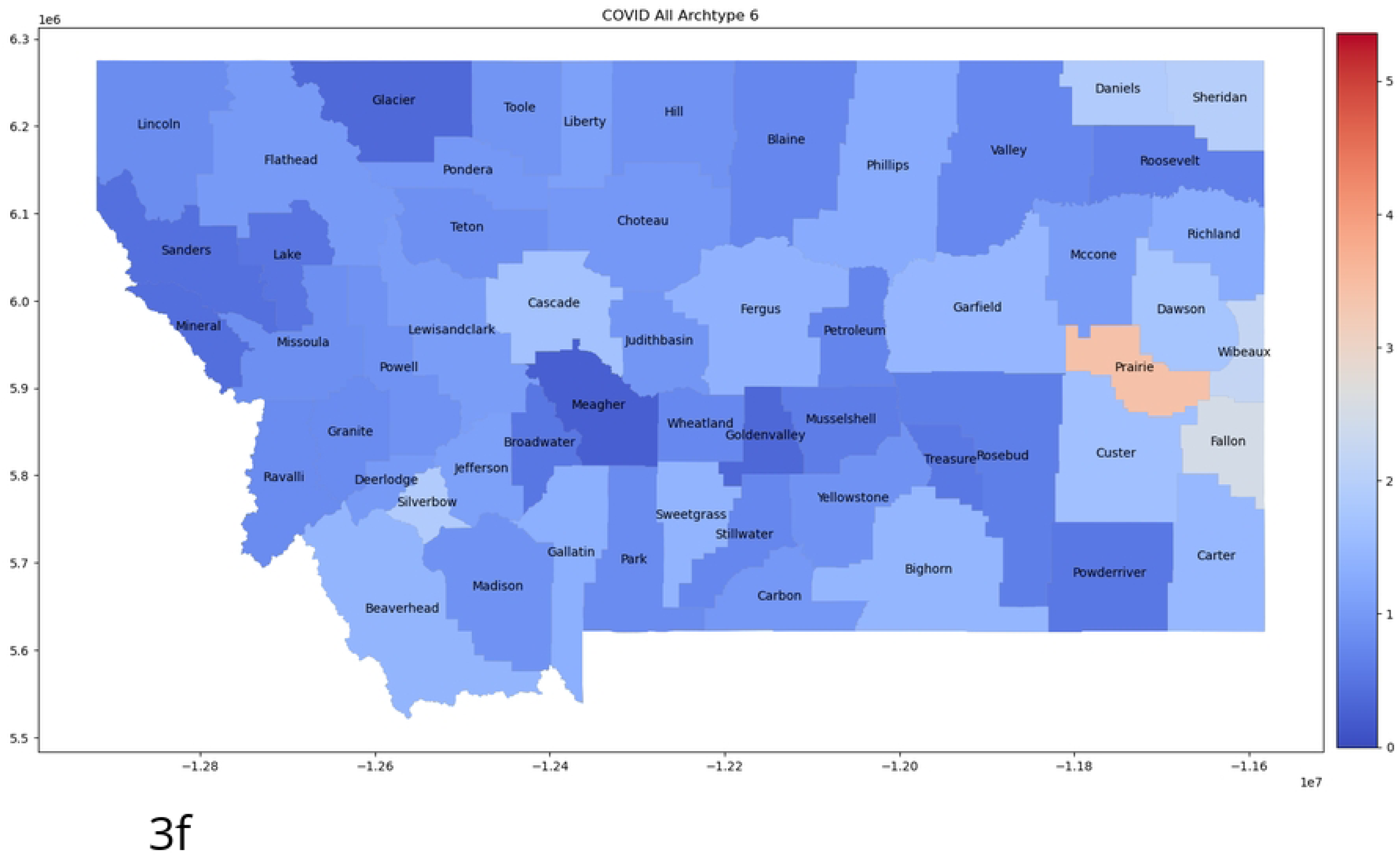
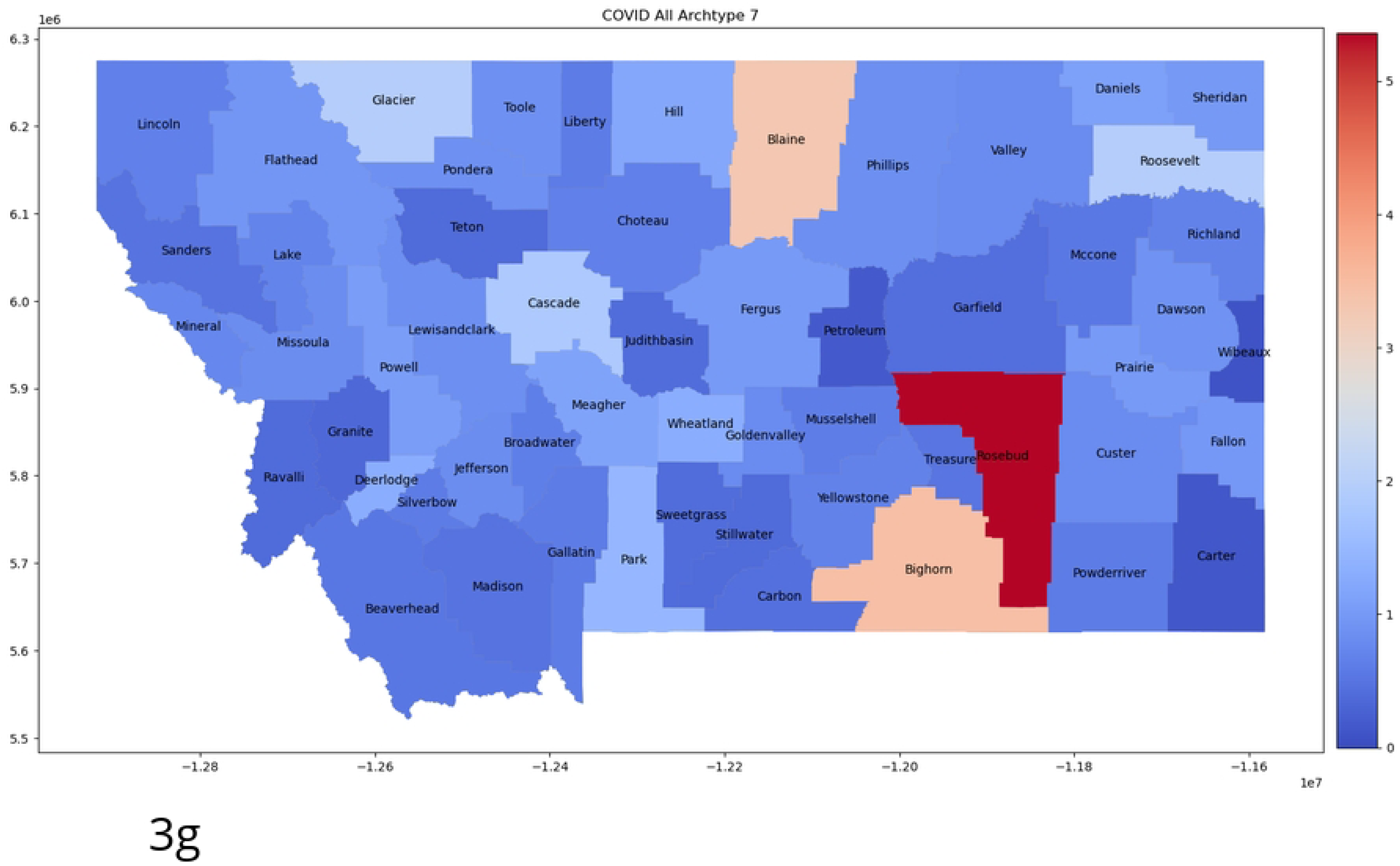
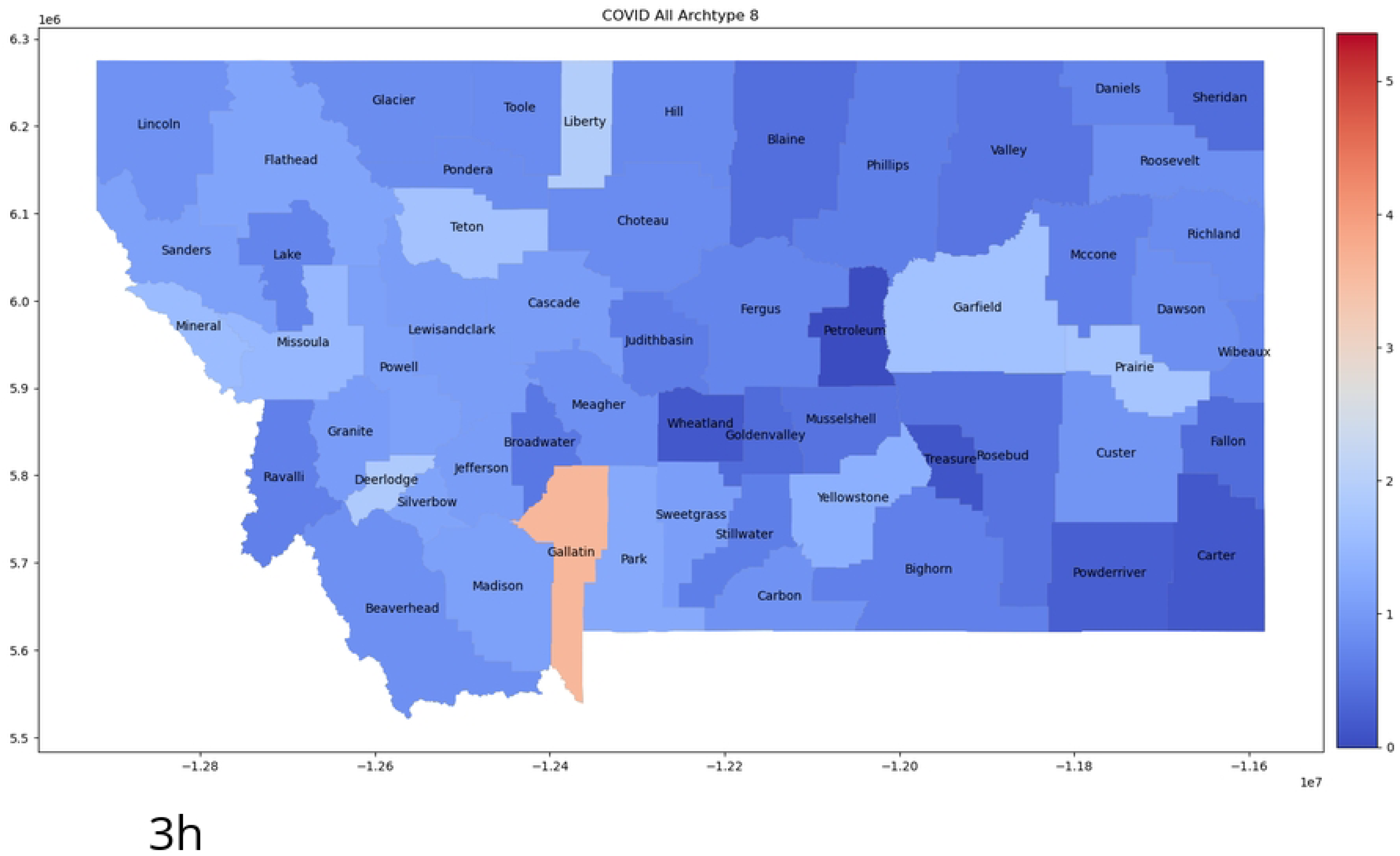
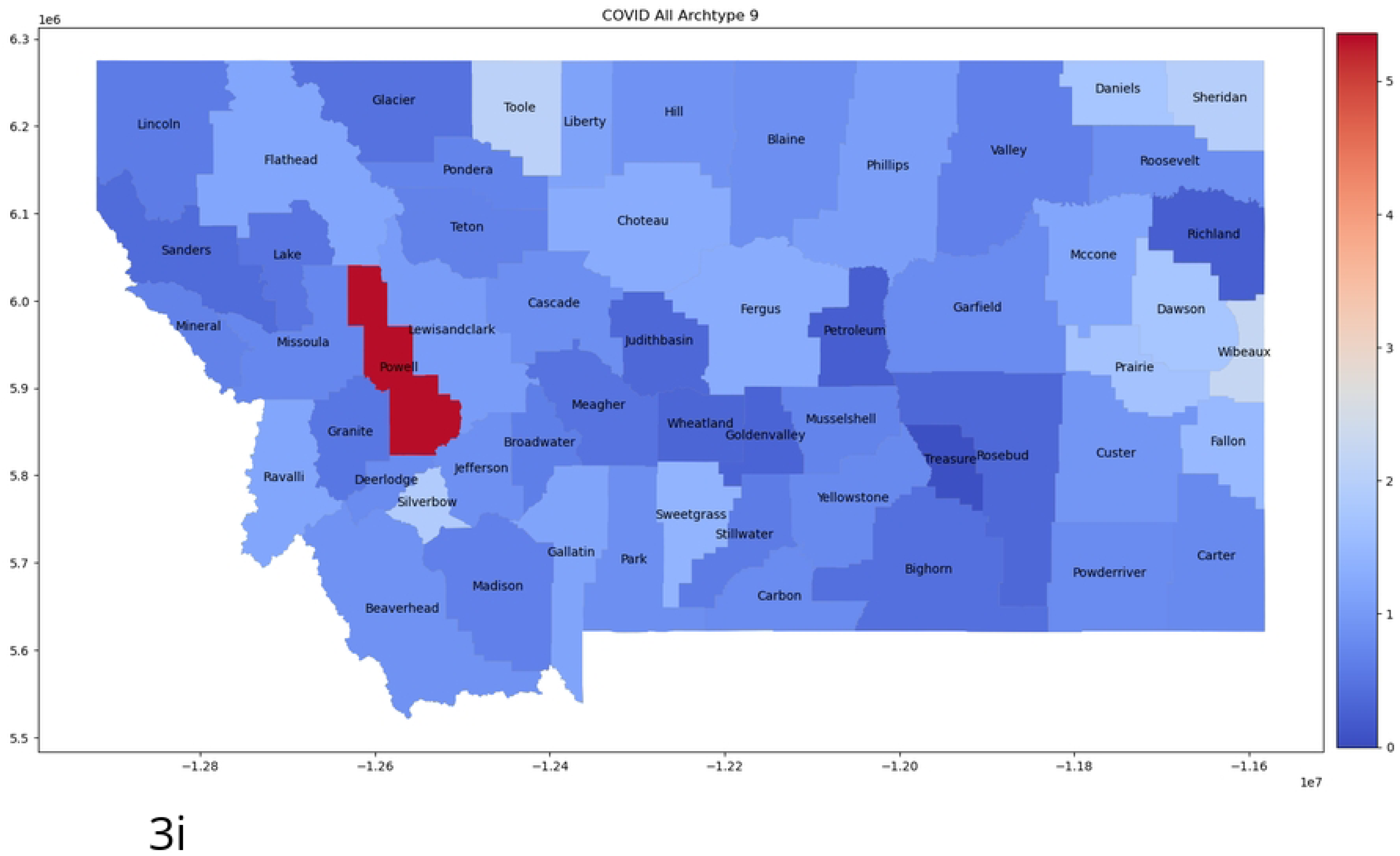
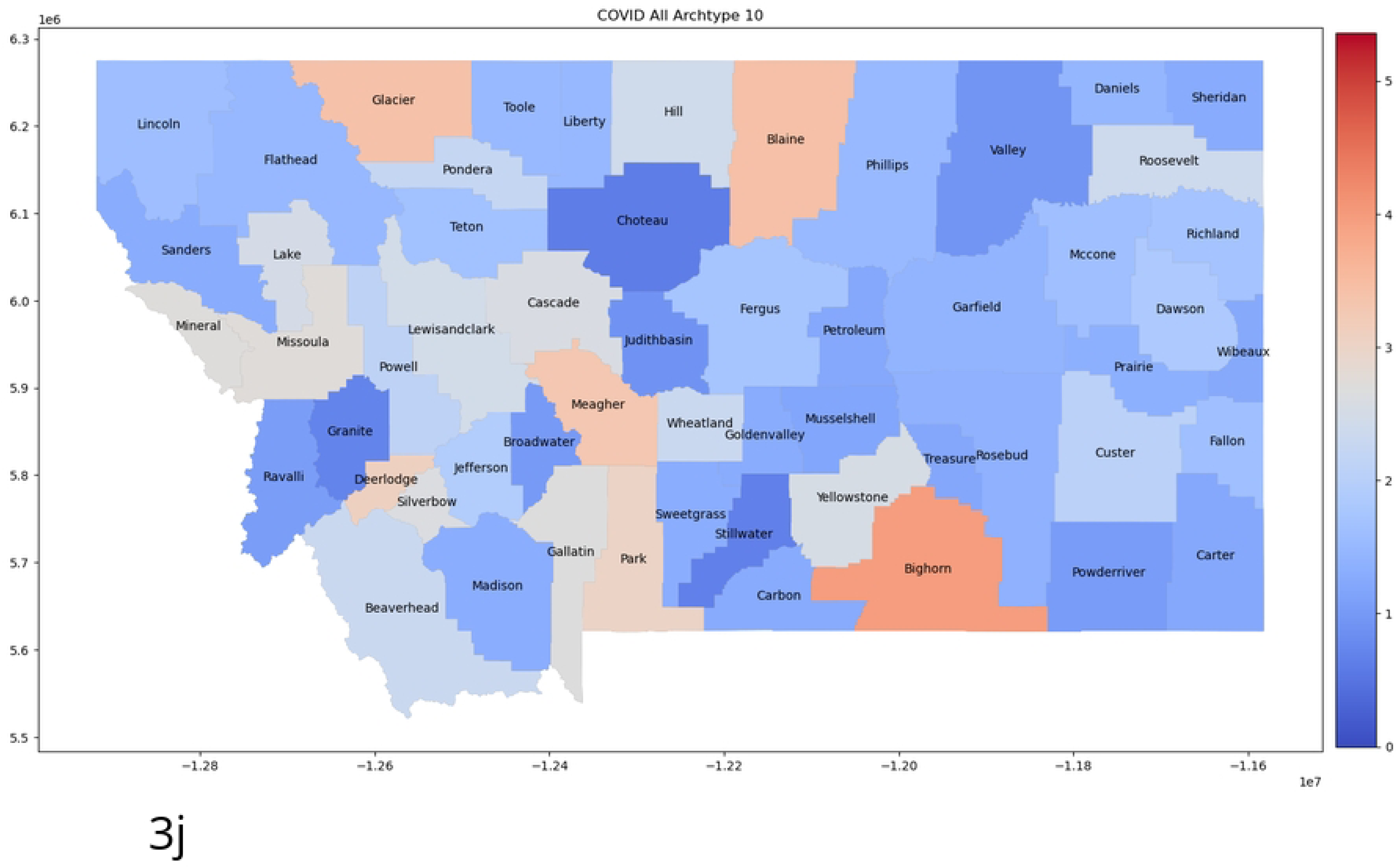
Ten Archetype set for all 56 counties. Presented as color-coded counties in the map of Montana. The first archetype [a] is nearly zero, as it captures the “no disease” state, and acts to “turn-off” the infection/spread in each county. The rest are in order: [b] archetype 2, [c] archetype 3, [d] archetype 4, [e] archetype 5, [f] archetype 6, [g] archetype 7, [h] archetype 8, [i] archetype 9, [j] archetype 10. Note that high *β* values are considered those greater than 1.5, mid-size between 0.75 and 1.5, and below 0.75 as low.

The archetype algorithm is strongly affected by outliers, as they will contribute largely to the RSS if not included included in the set. To remove these in a systematic way, we use techniques from information theory in the following section.

### Reducing Spatial Dimension and Removing Outliers with Information theory

As mentioned in the last section, normalizing the data by county population biases the size of the small population counties’ counts inordinately. Archetypal Analysis will choose such outliers in the data set to form archetypes, hence the small counties, with large per capita numbers, can drive the archetypes. In contrast, their time series are the most stochastic, and less likely to have any real predictive relationship with the other counties. To determine which counties should be included in the archetypal analysis, we computed the mutual information between all counties, and ranked counties according to their total mutual information (see also [4]).

Following the formulas in section 2, we first created histograms of the time series data for single counties, and joint histograms for each county with all the others. We note that the choice of bin size in these histograms will change the value of the entropy, but by choosing a fixed bin size for all the histograms, it is possible to compare the measures relative to each other.

Accordingly, we calculated the entropy with a uniform bin size and 30 partitions, for each county with respect to all others, creating a 56 by 56 matrix. To determine which counties have the highest MI in total, the MI row/column for each county is summed and ordered, to create the graph shown in Fig 4.

**Fig 4.**
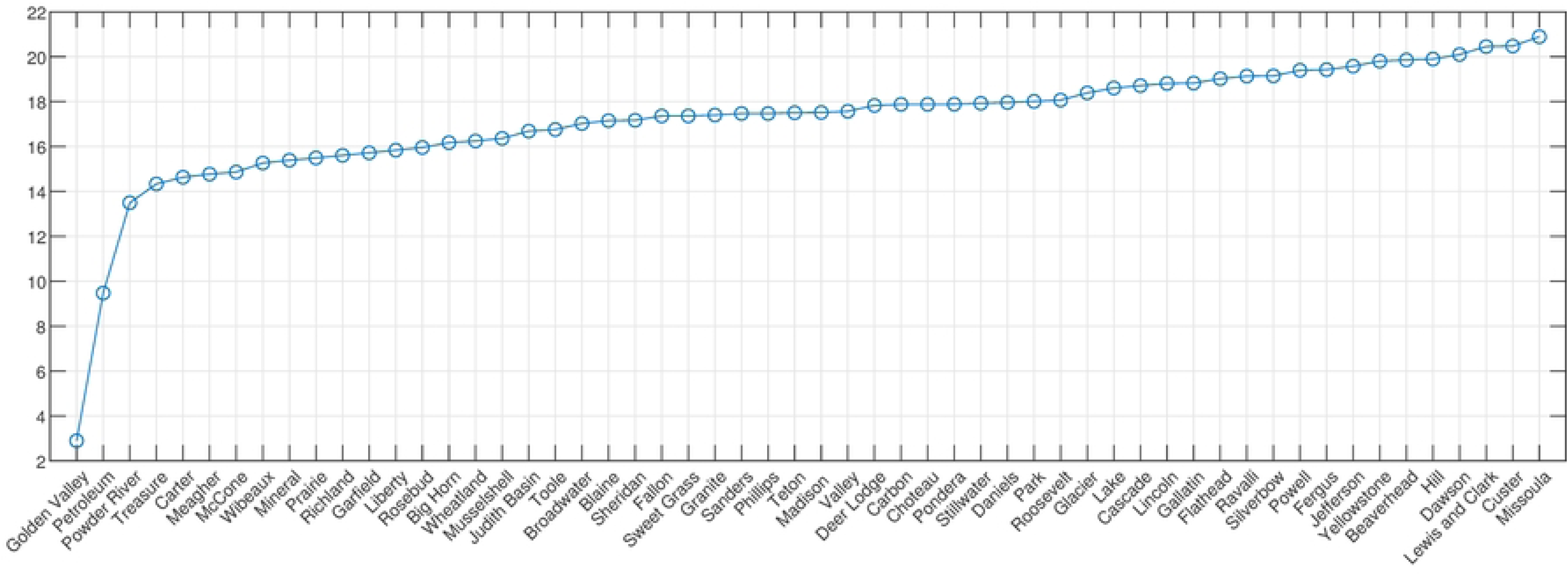
Total Mutual Information: all counties. Total mutual information (y-axis) across all counties in increasing order.

We choose the 17 top MI counties (see Fig.s 5 and 6) to analyze, as there is no clear elbow criterion except for removing the 2 very smallest population counties. Taking 17 includes all the large population center counties, and the smaller population counties that have larger MI. We also consider a smaller set of the the 10 largest population counties in the state in our analysis in the next section. Their populations, in alphabetical order, are: Cascade (81366), Flathead (103806), Gallatin (114434), Lake (30458), Lewis & Clark (69432), Lincoln (19980), Missoula (119600), Ravalli (43806), Silverbow (34915) and Yellowstone (161300). We note that the counties can be grouped into rough geographic subregions, where they are connected by state and/or Interstate highways, each with one major city or town. These are noted in Table 2.

**Fig 5.**
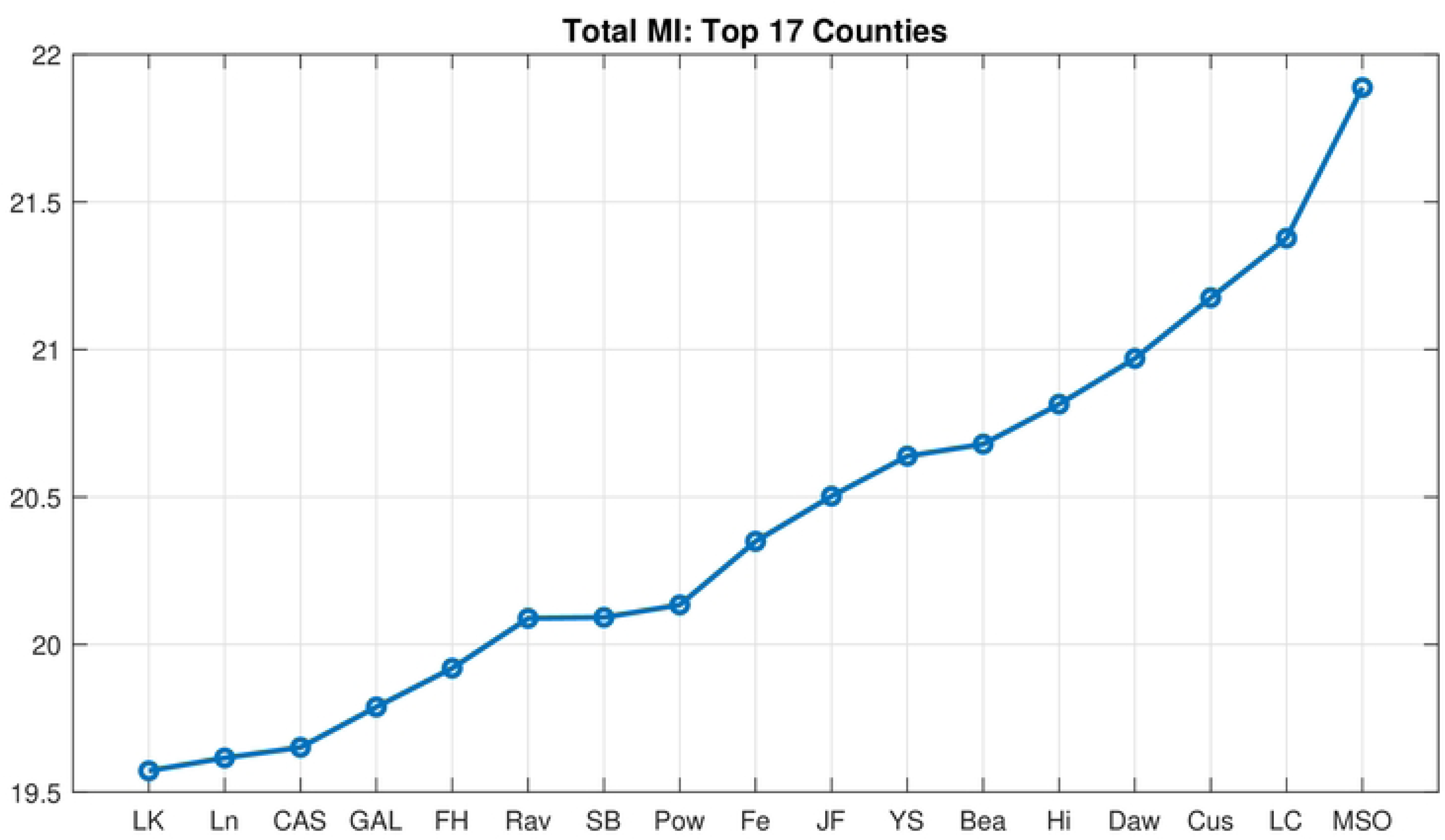
Total Mutual Information: top 17 counties. Total mutual information across top 17 MI counties in increasing order.

**Fig 6.**
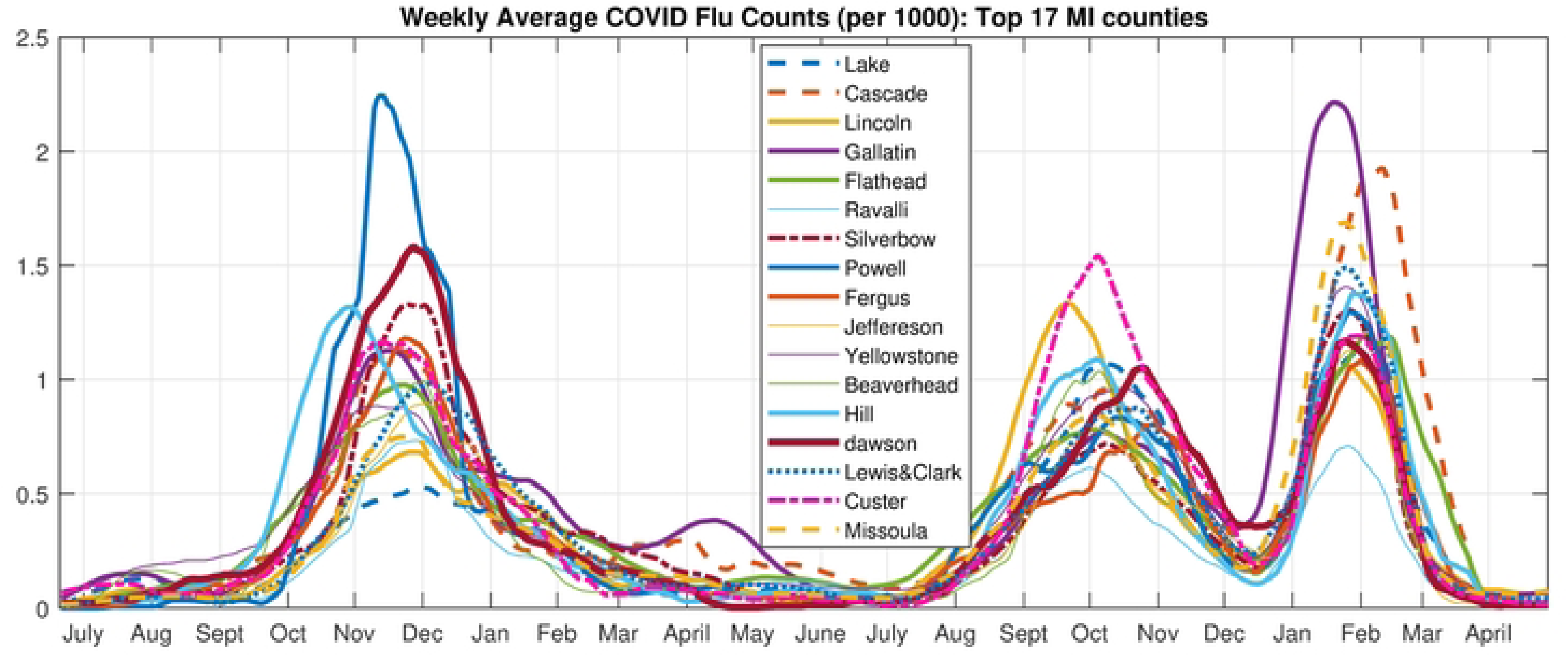
Time Series of Flu Counts. Case number time series for the 17 highest MI counties

**Table 1.** Archetype Composition-All County Set

| Archetype | Counties in Archetype |
| --- | --- |
| 1 | The zero or “no disease” archetype. |
| 2 | Widespread mid-size outbreak across state, largest in small population counties. |
| 3 | Isolated larger outbreak in Big Horn county, small in Phillips, very low in the rest. |
| 4 | Widespread low mid-size outbreak with a larger outbreaks in Fergus, Valley, and Musselshell counties, and slightly larger outbreaks in Park, Dawson, and Phillips counties. Note that these are all low population counties. |
| 5 | Very low level counts in all counties except for ten small population counties scattered all over the state, which have mid-size outbreaks. |
| 6 | Widespread mid-size outbreaks, with larger outbreaks in small population counties, with the exception of Silverbow. |
| 7 | Low to mid-size outbreaks in all counties except Big Horn, Cascade, Glacier, Blaine, Roosevelt, and Rosebud. The last three are very small population counties. |
| 8 | Large outbreaks in the high population counties of Yellowstone, Missoula and Gallatin, as well as the small population counties of Deer Lodge, Garfield, Liberty, Mineral, Prairie, and Teton. |
| 9 | Mid-size to low outbreaks in all counties except for nine very small counties which have very large outbreaks (Daniels, Dawson, Fallon, Powell, Sheridan, Silverbow, Sweet Grass, Toole, Wibaux) . |
| 10 | Widespread large outbreaks throughout state, both in large and small population counties. |

**Table 2.** Top Counties Grouped into Geographical Regions

| Region | Counties in Region |
| --- | --- |
| Northwest | Lincoln (Libby), Flathead (Kalispell), Lake (Polson) |
| North Central | Lewis & Clark (Helena, state capital), Cascade (Great Falls) |
| Southwest | Missoula (Missoula), Ravalli (Hamilton) |
| South | Silverbow (Butte), Gallatin (Bozeman), Yellowstone (Billings) |

### Archetypal Analysis of the maximal MI (17 County) Data Set

Archetypal analysis is used to decompose the COVID-19 counts into a limited number of spatial patterns over the set of the 17 high mutual information counties, and the reconstruction time series in terms of this set of patterns. Computing archetypal sets with increasing numbers gives Fig. 7, the residual sum of squares (RSS) for each set from 1 archetype to 20, and illustrates the drop-off in the error as the number of archetypes is increased. Note that the largest drop in RSS occurs after the first archetype, which, as mentioned earlier, captures the “no flu” state. Beyond that, the RSS declines more slowly to near zero as the number grows larger than about 15, as expected.

**Fig 7.**
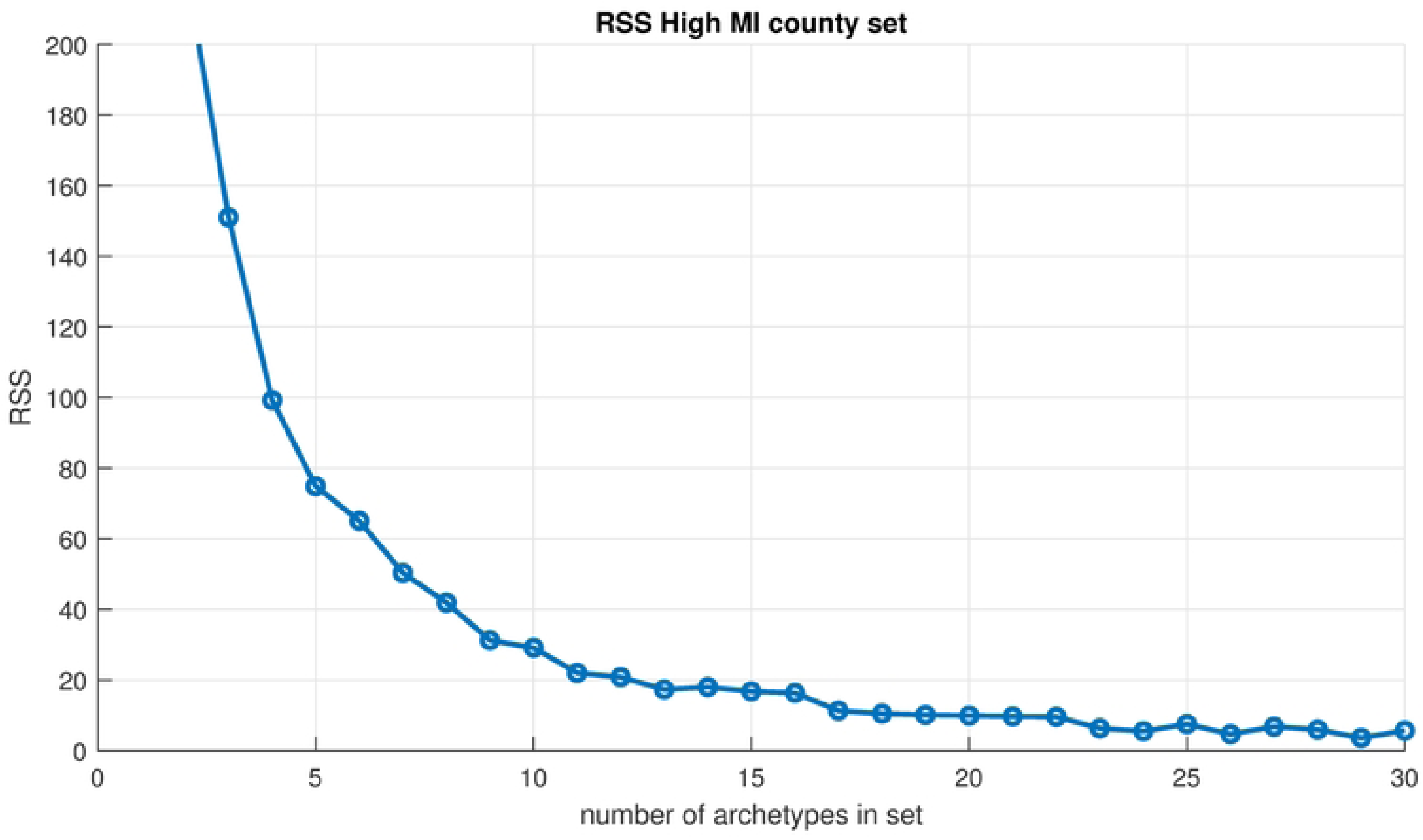
Scree plot. Residual sum of squares vs. number of archetypes being computed for the High MI County Set.

Using an elbow criterion on the scree plot to determine a cut-off, we choose 9 archetypes for the decomposition. We also examined an 8 archetype decomposition, and found that these 8 are included in the 9 archetype set. The additional archetype is archetype 2; a general widespread outbreak archetype, which should be included. Accordingly, we are presenting this 9 archetype set in our analysis here. See figure 8 for color maps of the archetypes. Each county is colored according to the *β* value it has for that archetype. Note that high *β* values are considered those greater than 1.5, mid-size are between 0.75 and 1.5, low are below 0.75. These values multiply the alpha values in the time series to give the data reconstruction. The 9 archetype set gives a good separation of the different outbreaks into distinct groups of archetypes, described below.

**Fig 8.**
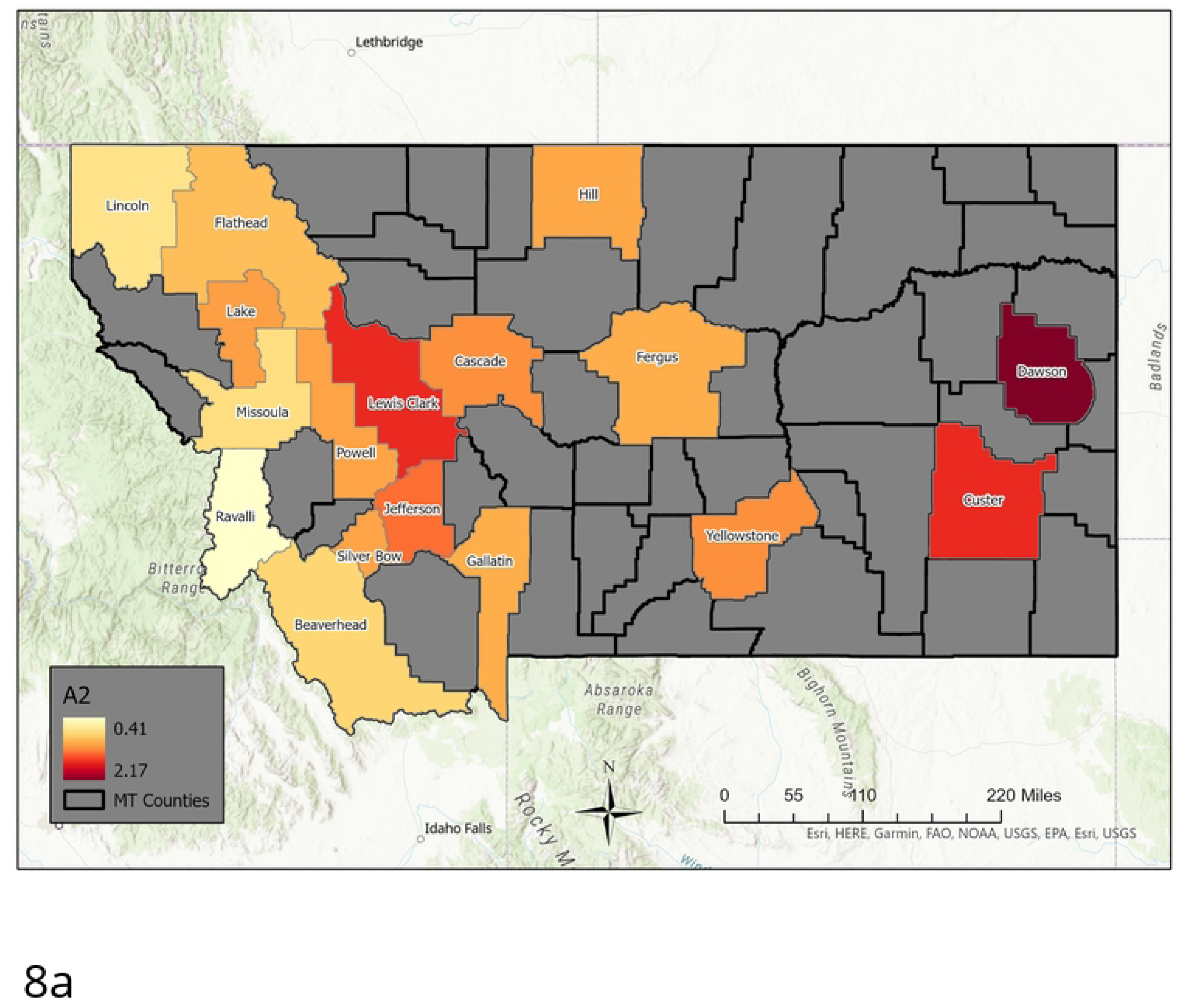
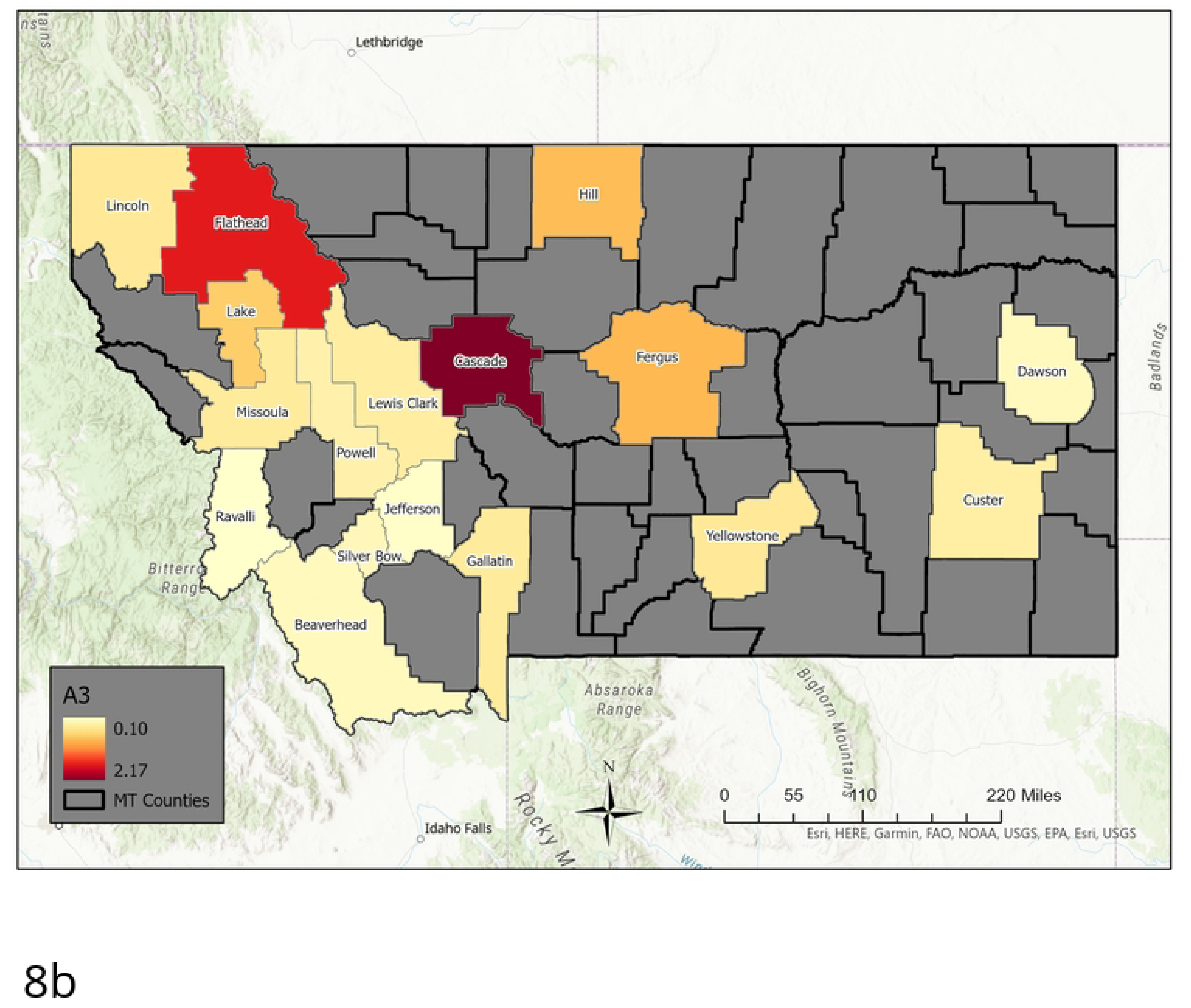
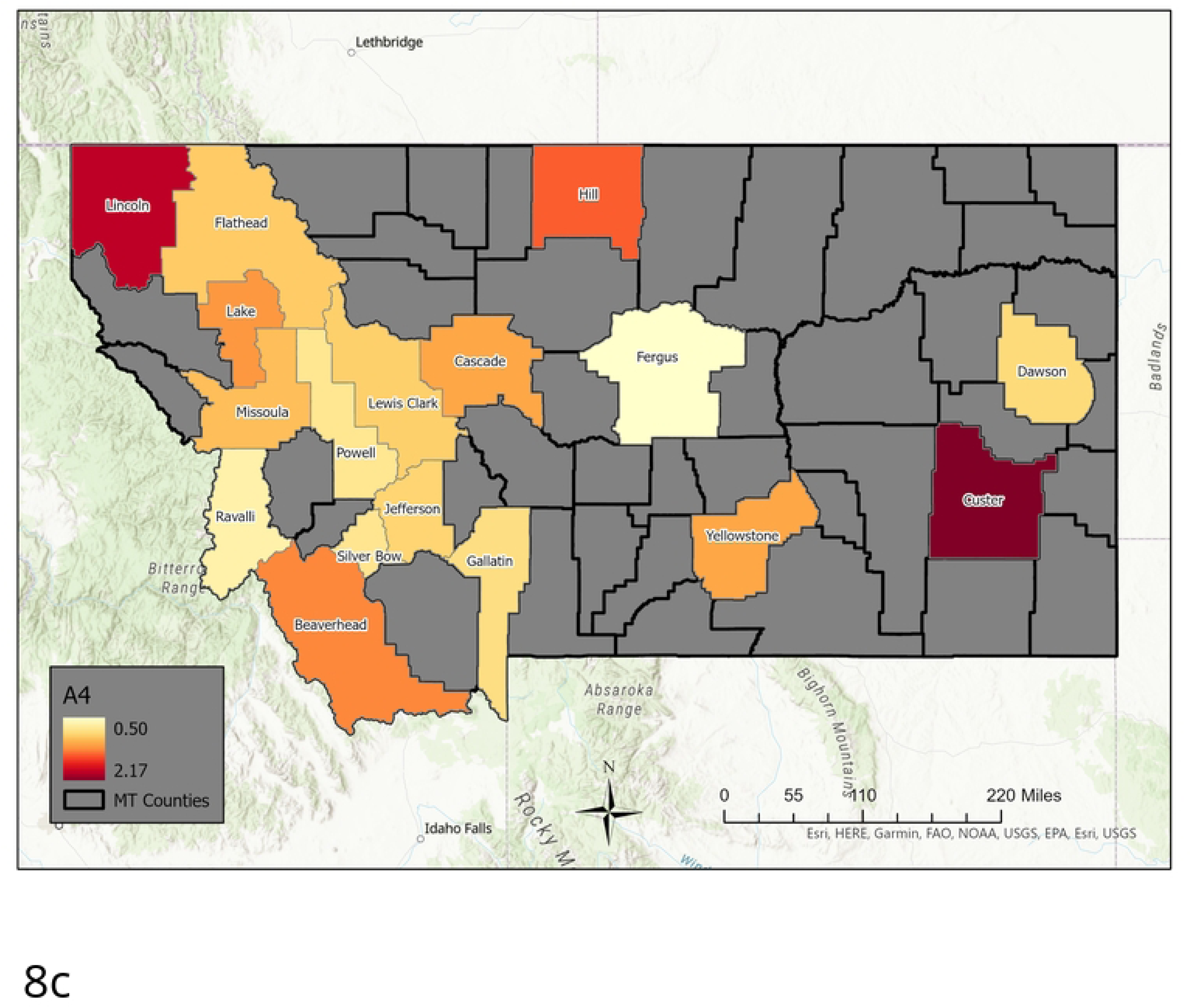
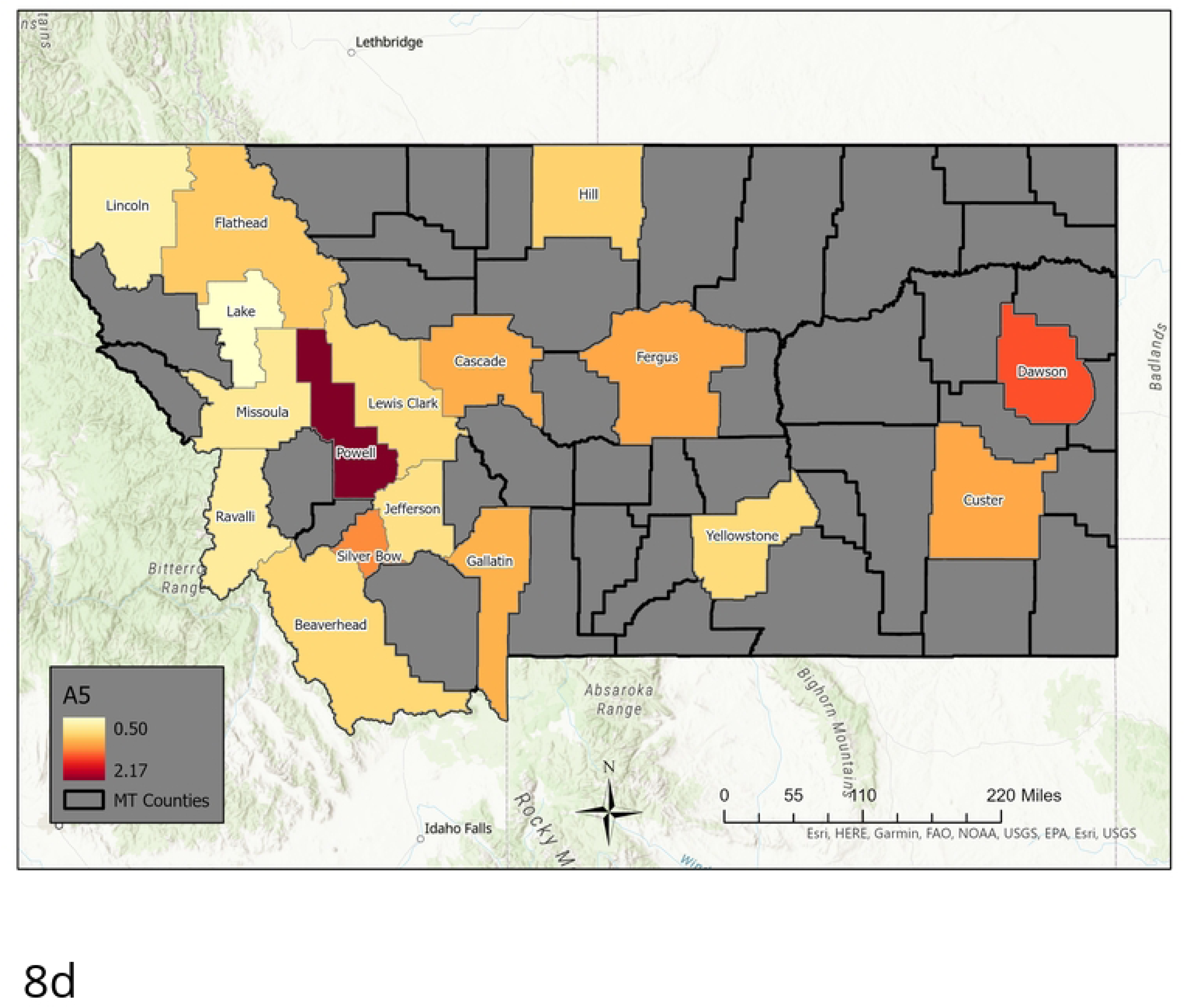
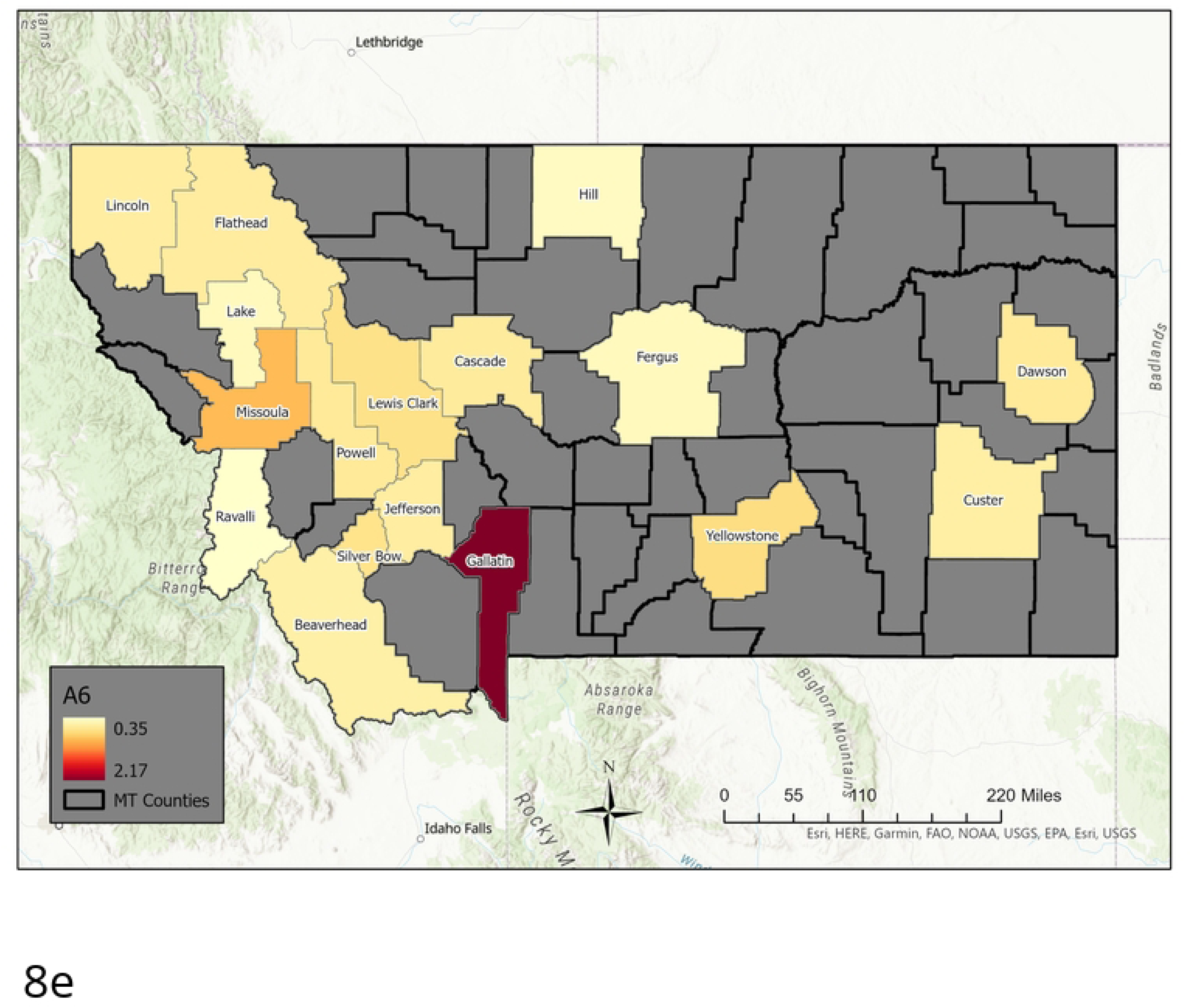
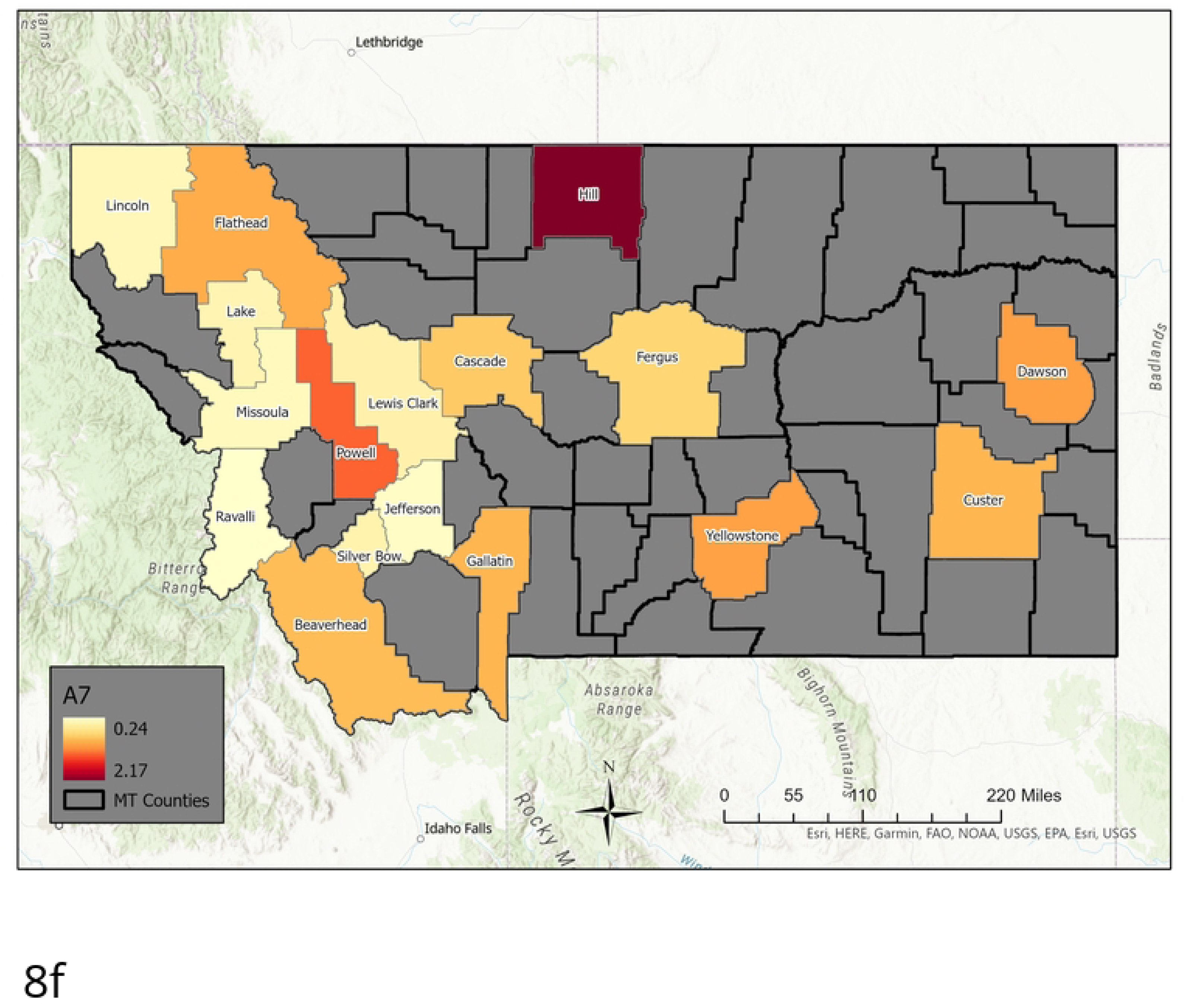
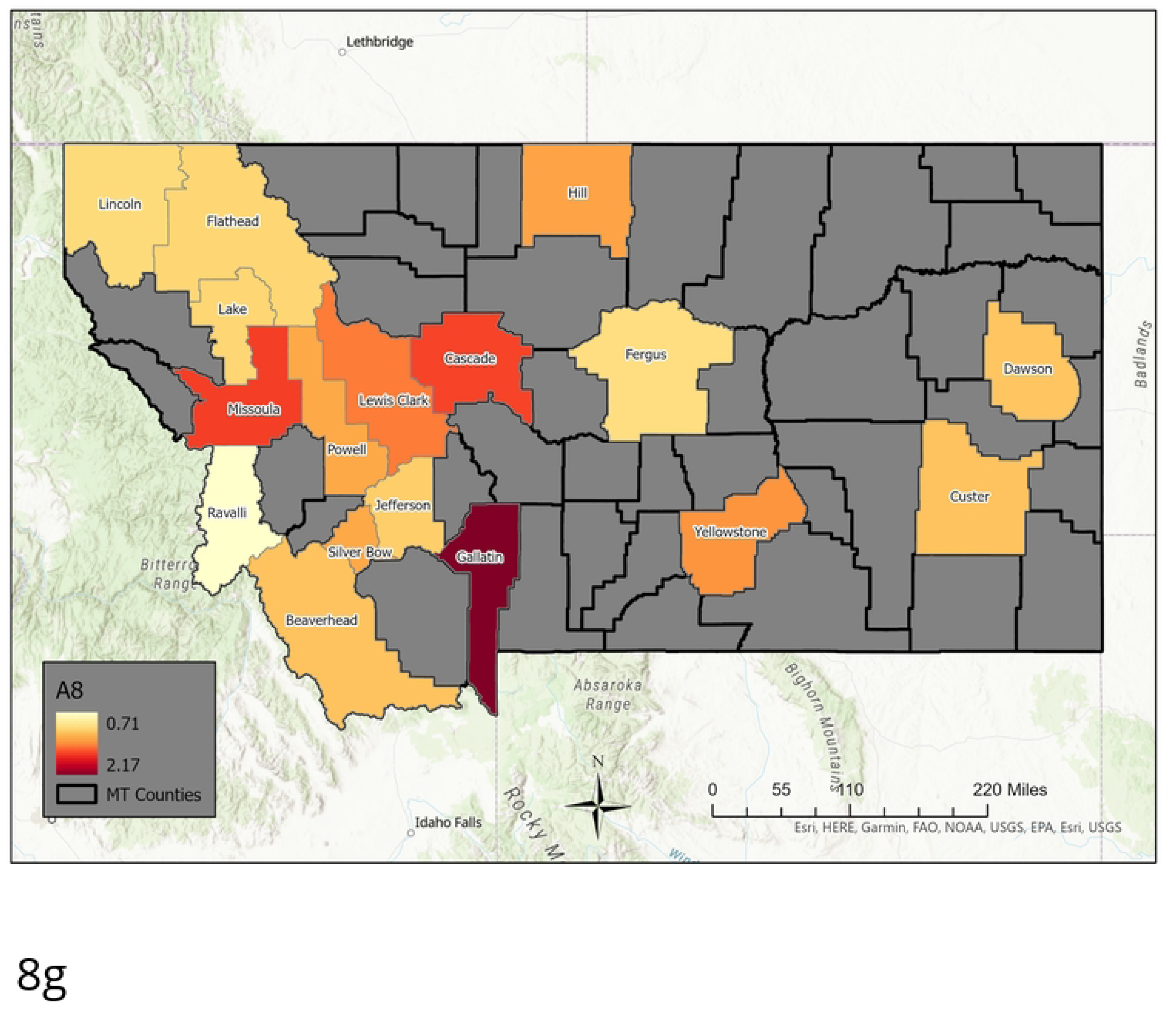
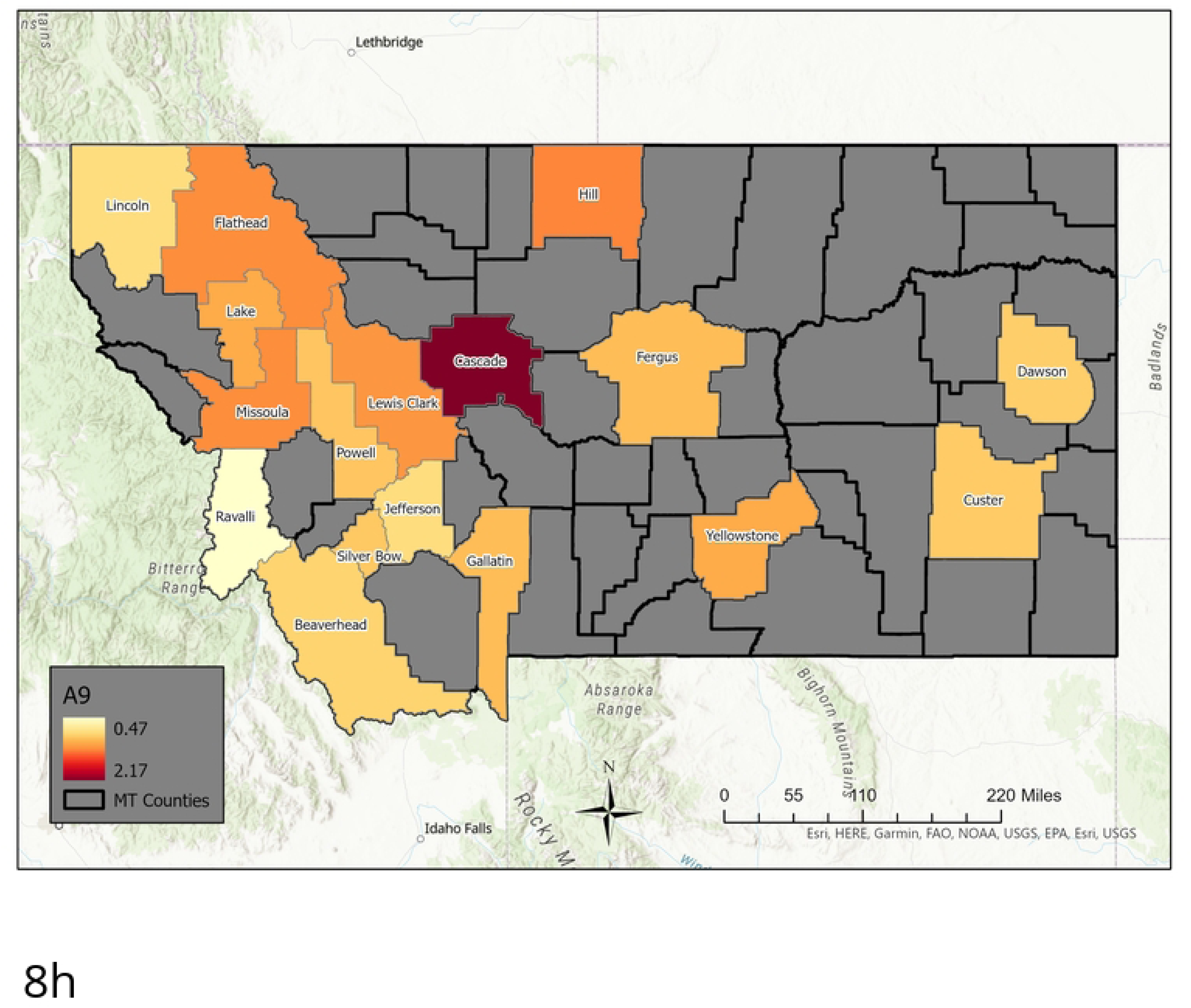
Nine Archetype set for the High MI County Set. Presented as color-coded counties in map of Montana. The first archetype is not included, as it captures the “no flu” state, and acts to “turn-off” the flu in each county. The rest are presented in order: [a] Archetype 2, [b] archetype 3, [c] archetype 4, [d] archetype 5, [e] archetype 6, [f] archetype 7, [g] archetype 8, [h] archetype 9. Note that high *β* values are considered those greater than 1.5, mid-size are between 0.75 and 1.5, low are below 0.75.

As the sum of the alpha time series is equal to 1 for each data point, it is fair to ignore one archetype; its alpha time series can be calculated from the remaining. We choose to ignore the zero archetype when analyzing in detail the anatomy and sequence of an outbreak, as it serves to turn the epidemic “off”. We further classify archetypes 1-9 in Table 3. Note that high *β* values are considered those greater than 1.5, mid-size are between 0.75 and 1.5, low are below 0.75. These values multiply the alpha values in the time series to give the data reconstruction.

**Table 3.** Archetype Composition-Maximal MI County Set

| Archetype | Counties in Archetype |
| --- | --- |
| 1 | The zero or “no disease” archetype. |
| 2 | Widespread low outbreak across state. |
| 3 | Widespread low outbreak with a concentration of a higher level of cases in Cascade and Flathead counties. |
| 4 | Mid-size over all, larger in Lincoln in the northwest, Hill in the north and Custer in the east. |
| 5 | Mid-size outbreaks overall, with larger outbreaks in Powell and Dawson. |
| 6 | Low level outbreaks in all counties except in Gallatin, which is high. |
| 7 | Mainly widespread outbreak, with lowest levels in Lincoln, Lake, Missoula, Ravalli, Lewis and Clark, Jefferson and Silverbow counties. Mid-size outbreaks in Flathead, Powell, Beaverhead, Cascade, Gallatin, Fergus, Yellowstone, Custer and Dawson. Larger outbreak in Hill. |
| 8 | Mid-size to very high level outbreaks over all. Largest in Gallatin, and large in Missoula, Lewis and Clark and Cascade counties. Powell and Silverbow have the next largest number of cases, then Yellowstone and Hill. The rest have more moderate size outbreaks. |
| 9 | Mid-size outbreaks in the contiguous region made up of Flathead, Lake, Missoula, and Lewis and Clark counties, with a large outbreak in adjacent Cascade county, and a mid-size outbreak in Hill county. The remaining counties have low mid-size outbreaks. |

The archetypes separate out clusters of counties which have simultaneous outbreaks (spatial), and the alpha’s give the sequence in which the outbreaks occur (temporal). The alpha time series indicate when each archetype is active in the time series. In Fig 9 we show how the 9 archetypes are present almost uniquely during the different outbreaks. The first outbreak is captured by archetype 7, 5 and 2, in that order. The Spring/Summer 2020 low outbreak is captured by archetypes 6 and 3, the Delta outbreak by 4 and 2, and finally the Omicron outbreak by 6, 8, 9 and 3. There are small contributions from other archetypes in each, e.g. archetype 2 is a low widespread outbreak, and is present at the end of the first outbreak and the Delta outbreak. Archetype 6 and 3 are present in the Spring/Summer outbreak, and to a lesser degree in the Omicron outbreak. These results suggest a different spatio-temporal spread during each outbreak.

**Fig 9.**
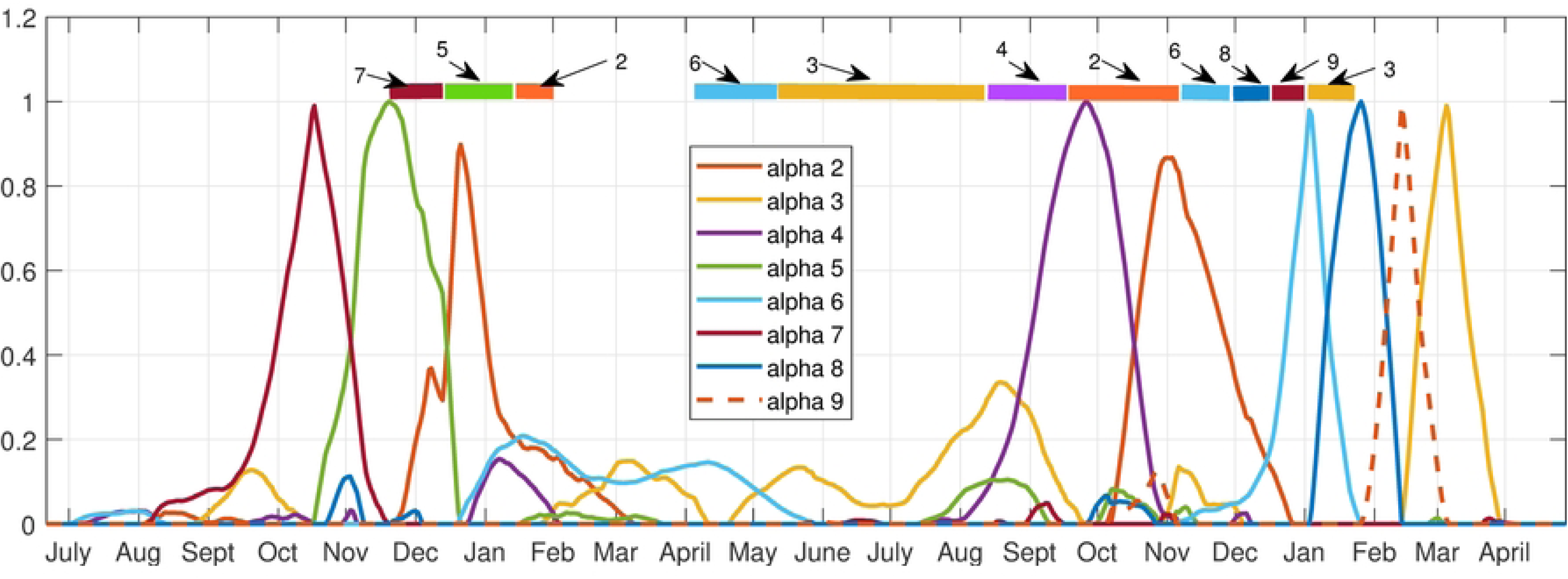
Alpha time series for the 9 archetype decomposition of the 17 county set. Bars across top are color-coded to show the dominant archetype during that time period.

The first outbreak begins with low counts except in Hill county (archetype 7), spreading to the central counties of Cascade and Fergus, along with Gallatin, whilst growing in the eastern counties of Custer and Dawson (archetype 5). It is largest in Powell, and mid-size in the rest of the western counties. The outbreak dies down to low-levels in all counties at the end (archetype 2). This outbreak thus spreads from the northern border of the state, to a mid-size outbreak in the central and eastern counties. The outbreak is mid-size in the west, with the exception of Powell, Silverbow and Gallatin, which are linked by I15 from Deer Lodge to Butte to Bozeman.

After that, in Spring/Summer archetype 6 shows declines in counts in all counties except Missoula and a large outbreak in Gallatin, then archetype 3 represents a further decline in counts in all counties except Cascade and Flathead, where a small increase occurs. The summer outbreak is mainly a sharp increase in cases in Gallatin county, represented again by archetype 6.

The Delta outbreak begins with largest counts in Lincoln (far northwest corner of the state) and Custer (far east side of the state), and mid-size elsewhere (archetype 4). Larger counts are also found in Hill, Yellowstone, Cascade, Lake and Beaverhead. The outbreak appears to spread in from the east and northwest. Then the transition to archetype 2 means a decline in all county counts except Dawson. Note that no other outbreaks use archetype 4.

The Omicron outbreak uses the most archetypes, beginning with a larger outbreak in Gallatin (archetype 6), to large counts in all counties, and a further increase of counts in Gallatin (archetype 8). Then a decline in Gallatin counts occurs with declines in all counties except Cascade (shown in archetype 9). It ends with low level outbreaks everywhere except Cascade and Flathead which are low mid-size (archetype 3). The outbreak follows a path from the initial outbreak in Gallatin county to a large outbreak in the central and western large population counties of Missoula, Lewis and Clark, Cascade, Powell and Silverbow, and east to Yellowstone. Then while these counties experience a decline in flu counts, there is a late surge in cases in Cascade, before all decay at the end of the outbreak.

This analysis is confirmed by comparing it with the time series of the counties during each outbreak, see Fig 10. This figure shows flu counts in the counties with significant contributions to different outbreaks, for comparison with the archetypal description above. We see that the flu initially appears in Hill county followed by Powell (represented by archetype 7), after which it spreads to a larger outbreak overall, with largest numbers in Powell, Silverbow and Dawson counties, represented by archetype 5. The last stage is characterized by large counts in Gallatin county, mid-size in Silverbow, Yellowstone, Lewis and Clark and Missoula counties, and smaller in the rest (seen in archetype 2). The smaller outbreak in summer 2020 is captured by archetype 6 and 3, with largest counts in Gallatin county followed by larger counts in Cascade and Flathead counties. The Delta variant outbreak begins in September, with archetype 4, switching to archetype 2. Archetype 4 is large in Lincoln, Custer, Hill and Beaverhead counties, and 2 is a widespread outbreak which is largest in Dawson county. We see in the count time series that Lincoln and Custer do dominate initially, with Hill and Beaverhead also larger. As Lincoln and Custer counts decline, Dawson emerges. The Omicron outbreak begins in December with archetype 6, switching to 8, then 9, then 3. This reflects the initial large case numbers in Gallatin county, followed by widespread outbreak with large counts also in Missoula, Cascade, and Lewis and Clark counties, then to large counts in Cascade (archetype 9) and finally decaying in all counties except Cascade and Flathead (archetype 3).

**Fig 10.**
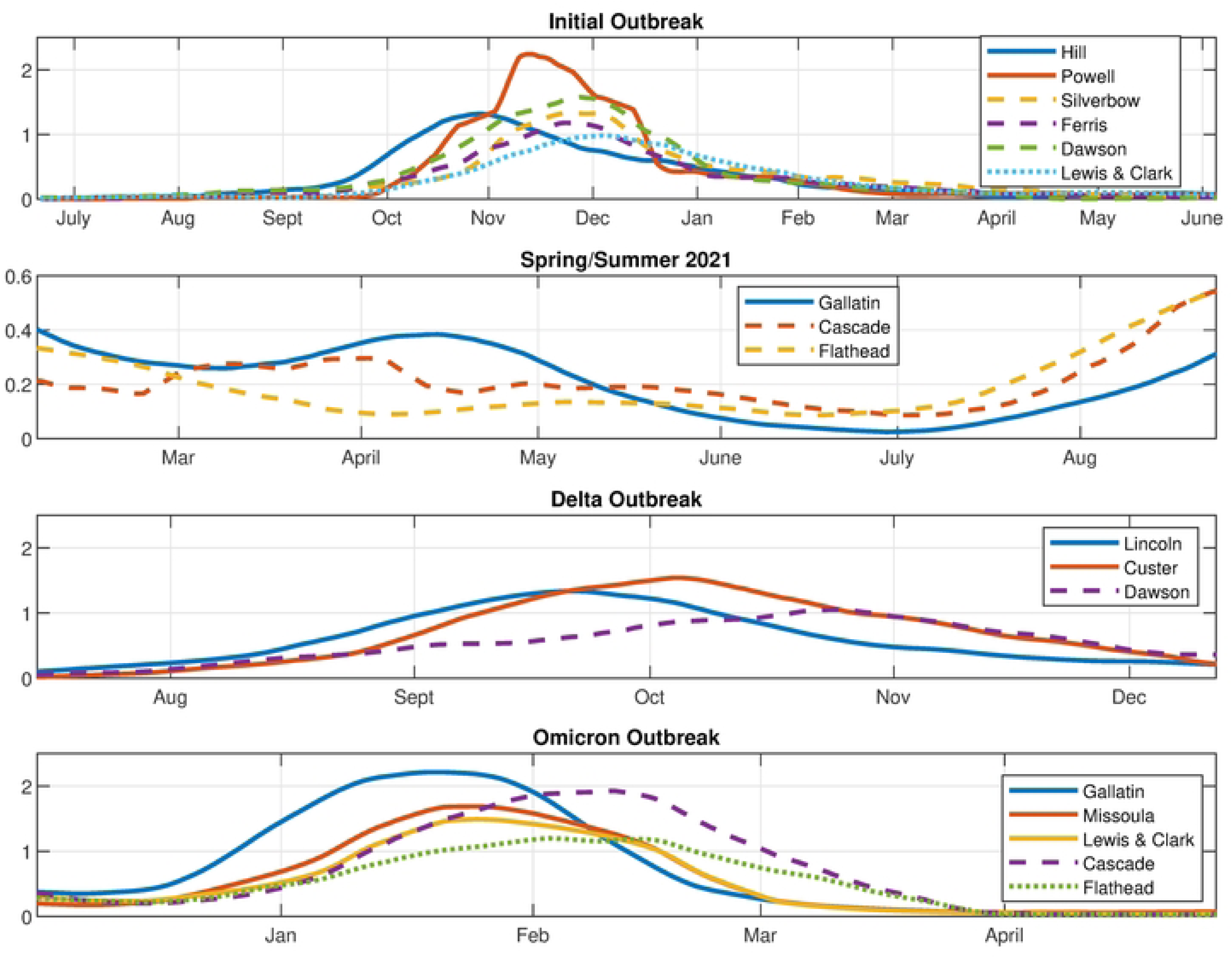
Time series of flu counts. Counties with significant contributions to the different outbreaks: Initial, Spring/Summer, Delta and Omicron (from the 17 county data set).

### Archetypal Analysis of High Population Center Set

We next consider the counties with large population centers. In decreasing order of population they are: Yellowstone (161300), Missoula (119600), Gallatin (114434), Flathead (103806), Cascade (81366), Lewis and Clark (69432), Ravalli (43806), Silverbow (34915), and Lake (30458), Lincoln (19980). The time series for each is plotted in Fig 11.

**Fig 11.**
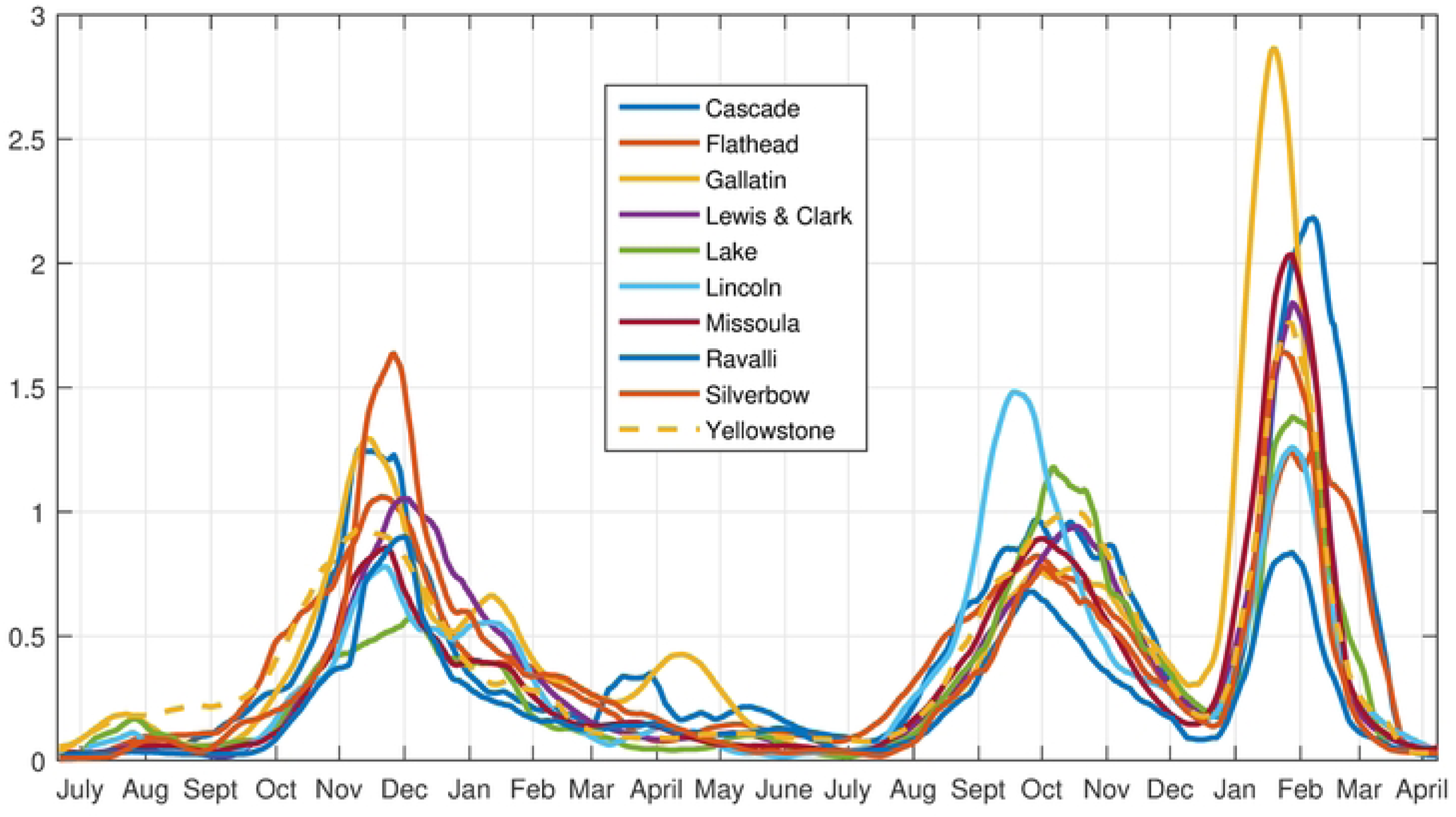
Time Series for the Large Population County Set.

Fig 12 shows the residual sum of squares (RSS) for each truncation from 1 archetype to 20. For this analysis we choose a truncation to 6 archetypes, which captures roughly 98% of the variance. Thus, we have reduced the dimension of the data set from 10 to 6, or really 5, because the alpha time series must sum to one at each time point.

**Fig 12.**
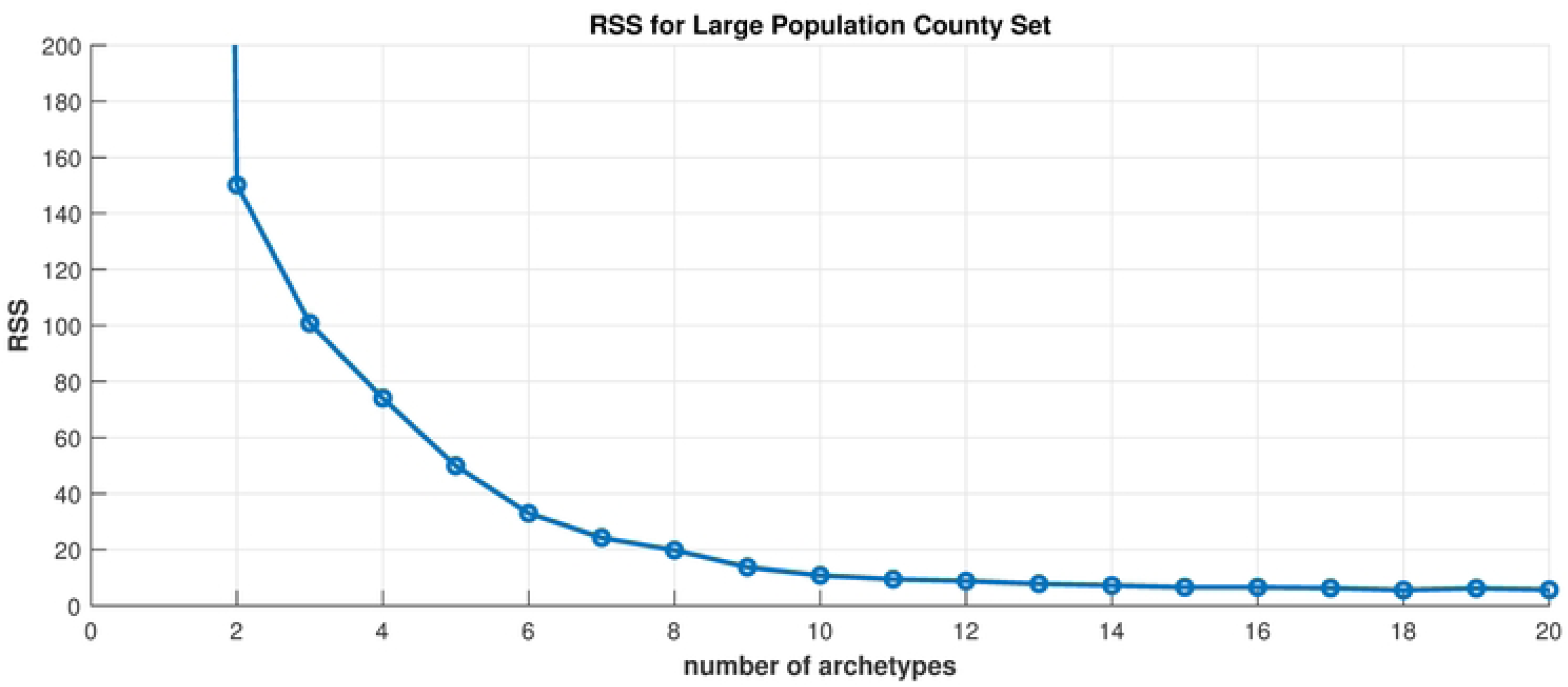
Scree plot. Residual sum of squares vs. number of archetypes being computed for the Large Population County Set.

In Figure 13 we show color maps of each archetype. Again, the archetypes are ordered according to the overall size of their contribution to the reconstruction. The alpha time series (Figure 14) show when each archetype is active in the time series. We classify archetypes 1-6 in Table 4.

**Fig 13.**
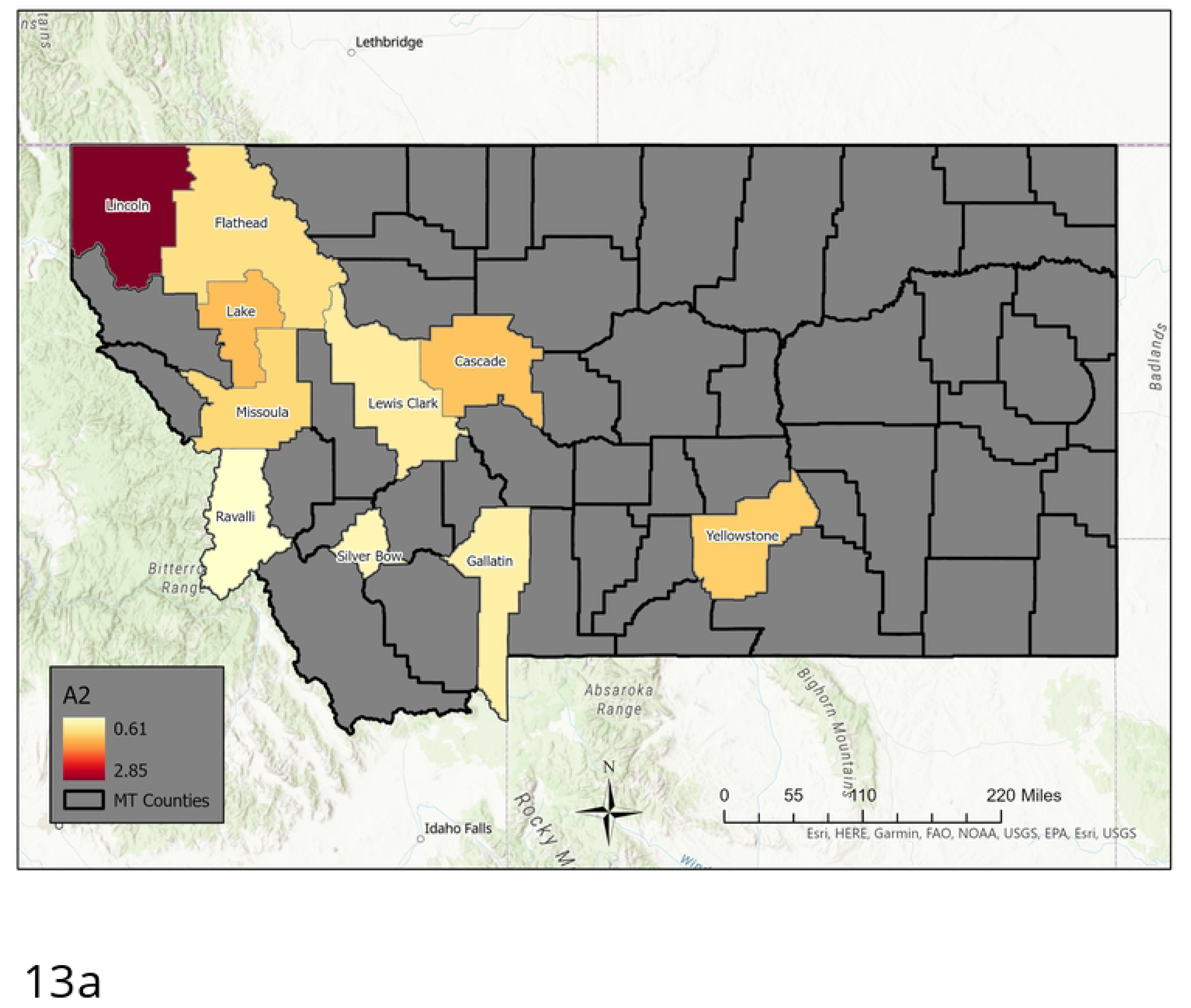
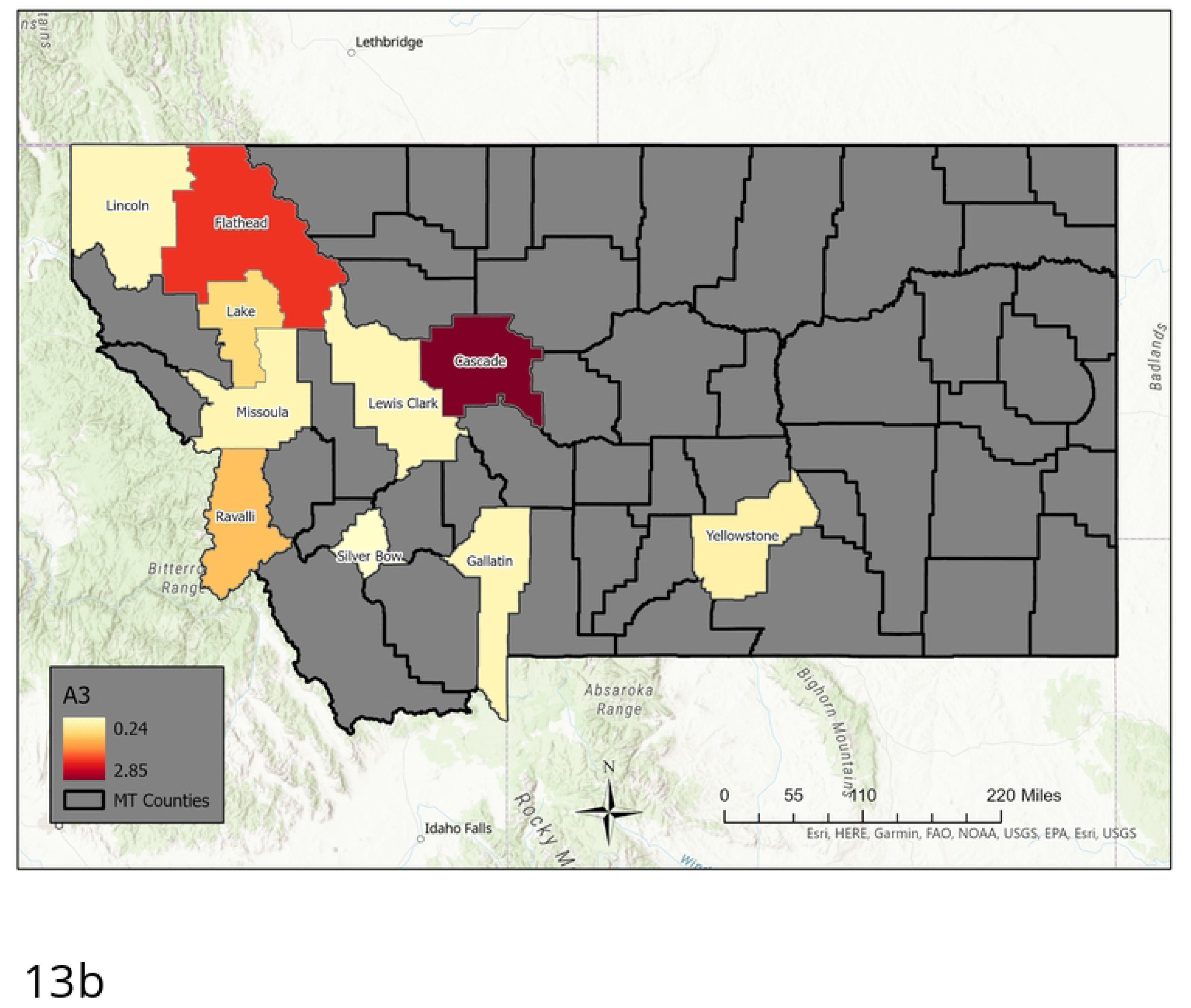
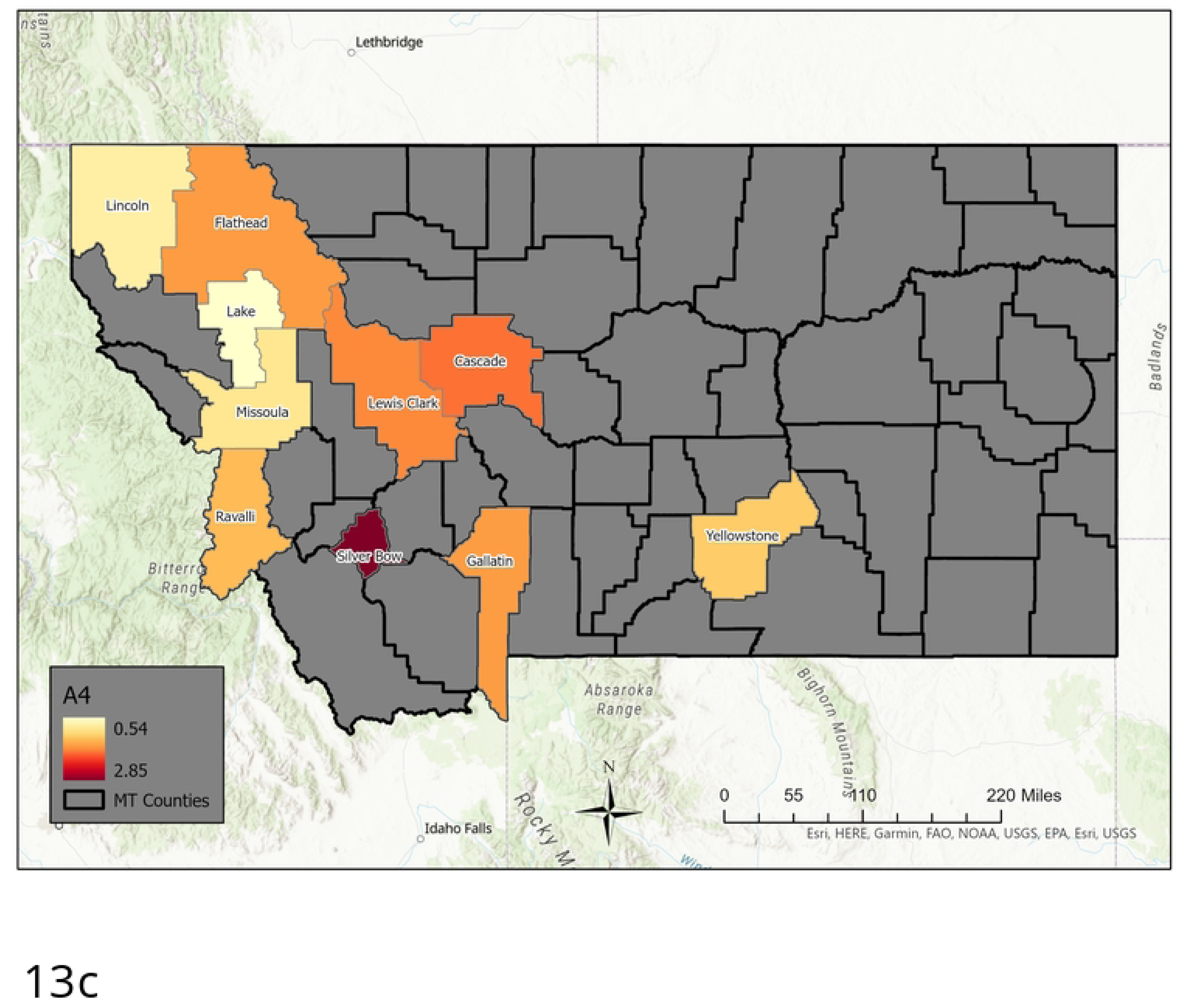
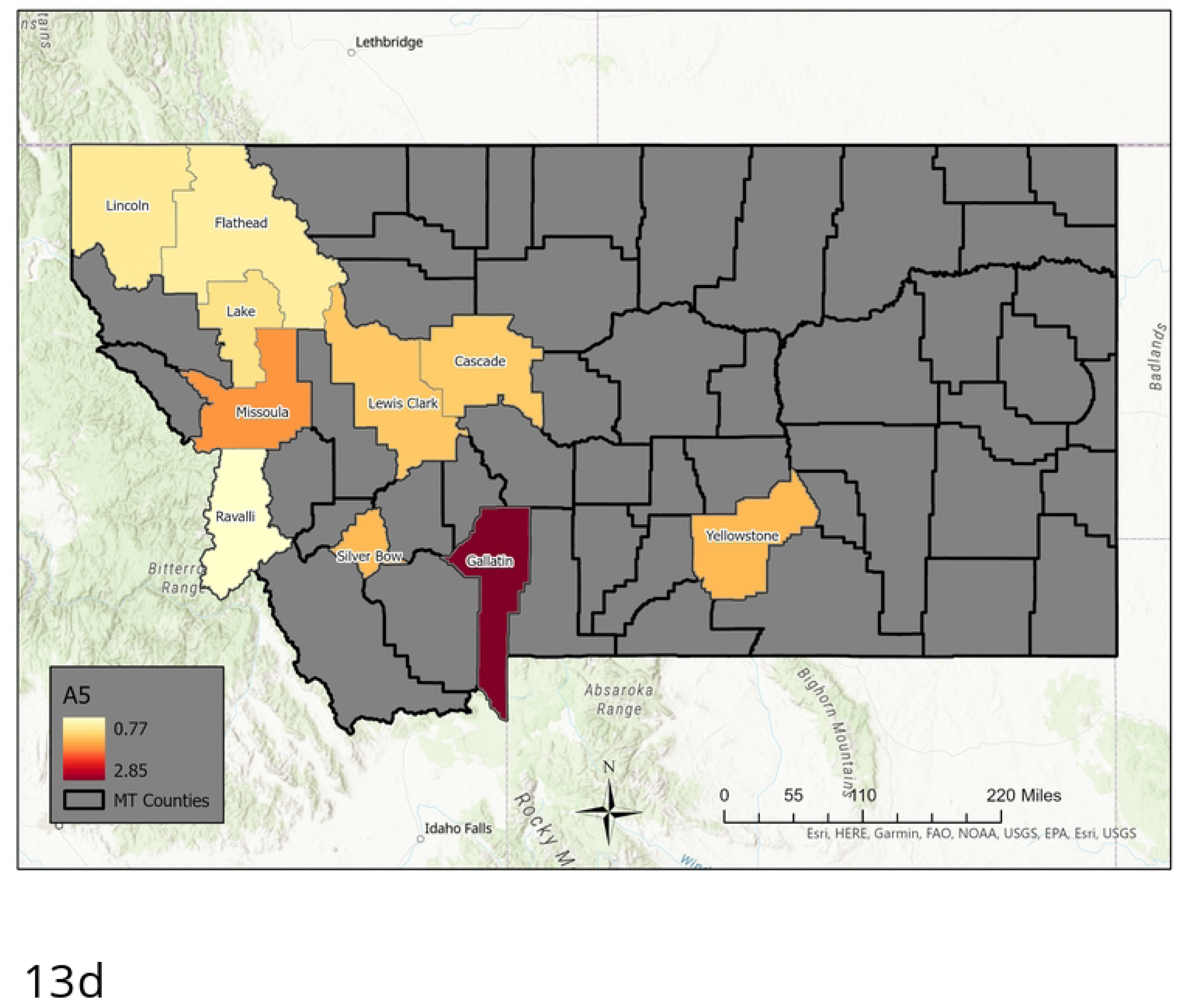
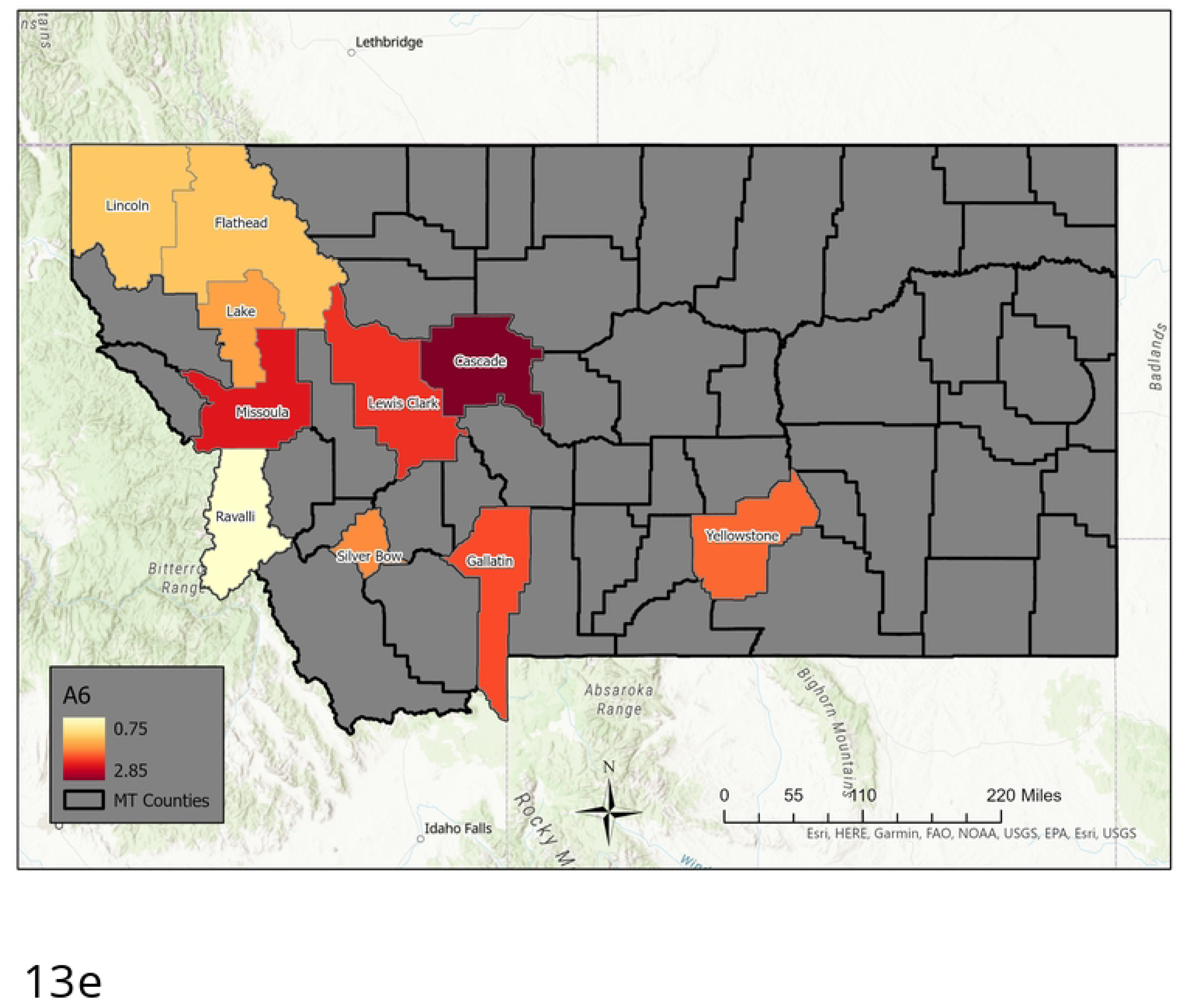
Six Archetype set for the Large Population County Set. Presented as color coded counties in Montana. Again, the first archetype is not included, as it captures the “no disease” state, and acts to “turn-off” the flu in each county. The rest are in order: [a] archetype 2, [b] archetype 3, [c] archetype 4, [d] archetype 5, [e] archetype 6.

**Fig 14.**
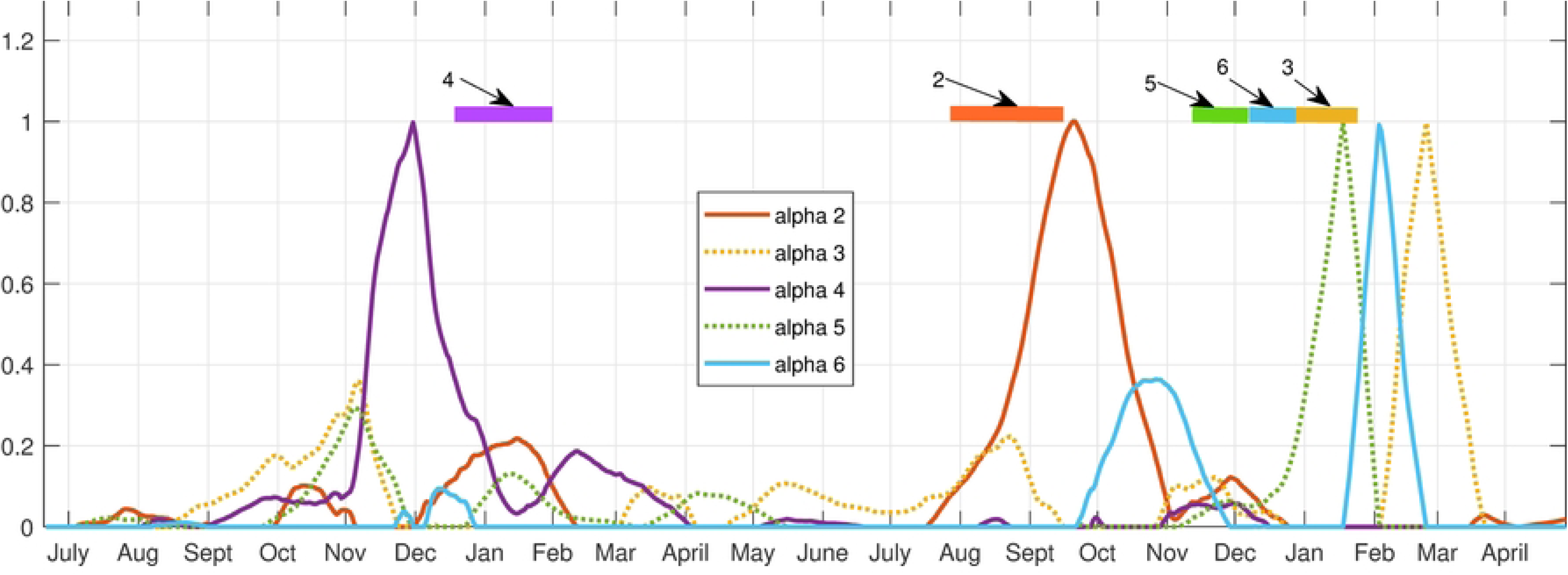
Alpha time series. The 6 archetype decomposition of the 10 county set. Bars across top are color coded to show the dominant archetype during that time period.

**Table 4.**
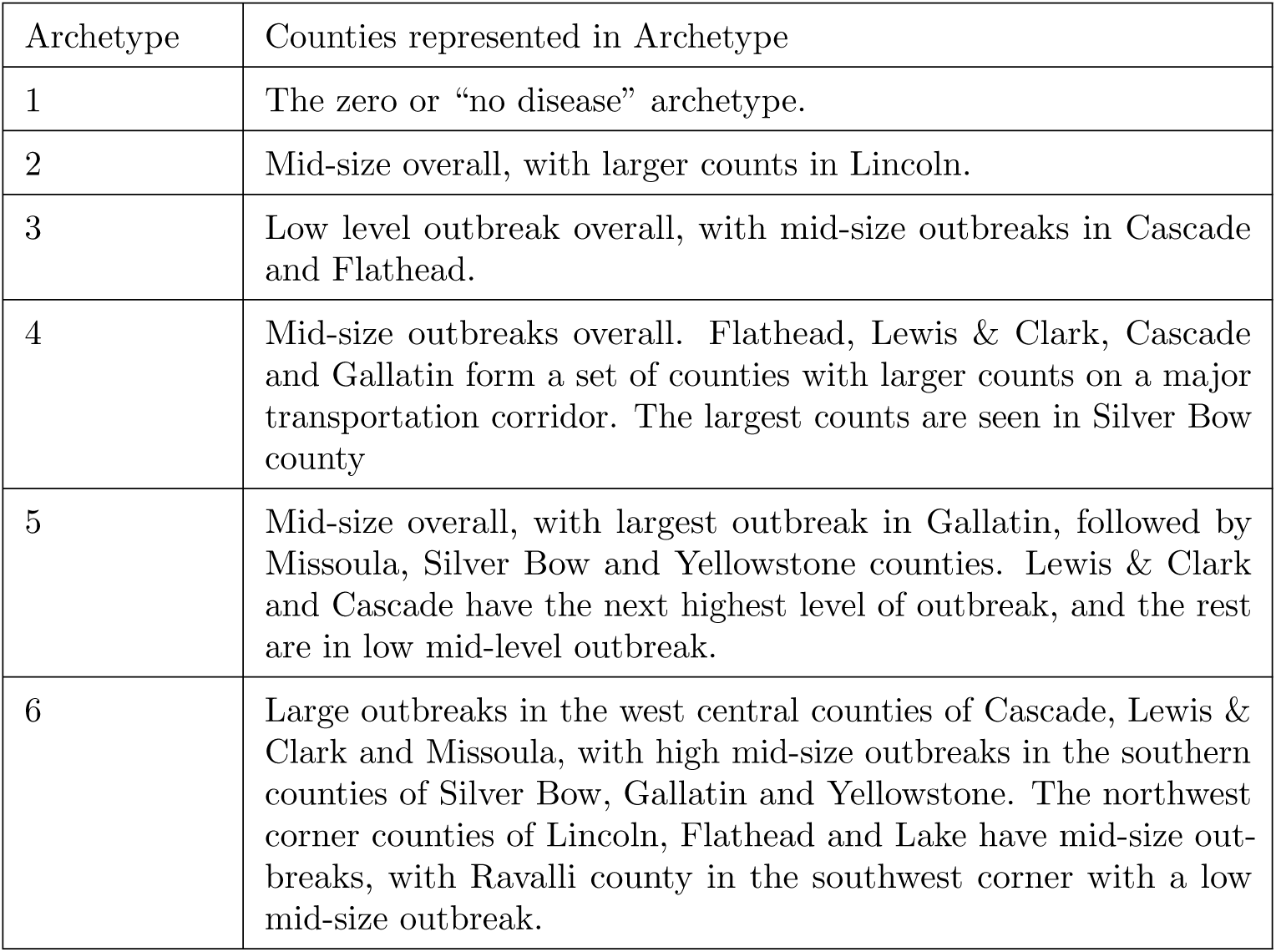
Archetype Composition-Large Population County Network

| Archetype | Counties represented in Archetype |
| --- | --- |
| 1 | The zero or “no disease” archetype. |
| 2 | Mid-size overall, with larger counts in Lincoln. |
| 3 | Low level outbreak overall, with mid-size outbreaks in Cascade and Flathead. |
| 4 | Mid-size outbreaks overall. Flathead, Lewis & Clark, Cascade and Gallatin form a set of counties with larger counts on a major transportation corridor. The largest counts are seen in Silver Bow county |
| 5 | Mid-size overall, with largest outbreak in Gallatin, followed by Missoula, Silver Bow and Yellowstone counties. Lewis & Clark and Cascade have the next highest level of outbreak, and the rest are in low mid-level outbreak. |
| 6 | Large outbreaks in the west central counties of Cascade, Lewis & Clark and Missoula, with high mid-size outbreaks in the southern counties of Silver Bow, Gallatin and Yellowstone. The northwest corner counties of Lincoln, Flathead and Lake have mid-size outbreaks, with Ravalli county in the southwest corner with a low mid-size outbreak. |

As in the largest total MI data set, each archetype is almost uniquely identified with one of the major outbreaks. Archetype 4 (Cascade, Flathead, Gallatin, Lewis & Clark, Silverbow) peaks during the first outbreak, archetype 2 (Cascade, Lake, Lewis & Clark, and Yellowstone) during the Delta outbreak. Archetypes 5 (Gallatin, Missoula), 6 (Cascade, Missoula, Lewis and Clark) and 3 (Cascade and Flathead) illustrate 3 different phases in the Omicron outbreak. These archetypes also make contributions to the shoulders of the major outbreaks, and 3 and 5, at low levels, are used to represent the summer 2021 isolated low level outbreak.

In the initial outbreak Gallatin, Cascade and Flathead counts rise first, which are contained in archetype 3, then replaced by archetype 4, which reflects a widespread outbreak with a larger component in Silver Bow county in late November 2020. This is the dominant archetype for the initial outbreak. The declining outbreak in January is captured by archetype 2 and 5 (widespread lower level outbreak, with a larger component in Gallatin) followed by archetype 4 again in February (fig 15 a) The spring/summer persistent outbreak is represented by low level archetype 5 switching to low level archetype 3, giving a rise in Cascade county followed by a rise in Gallatin county, with low level outbreak in other counties (see fig 15 b)). The fall 2021 Delta outbreak is initiated with low alpha 3 transitioning to large alpha 2, as it begins in Cascade and Flathead counties, then spreading to all counties with a larger component in Lincoln. Archetype 2 is thus the dominant archetype for the Delta outbreak. It ends as alpha 2 declines and alpha 6 rises, reflecting the decline in Lincoln counts and the growing counts in all other counties, especially the counties with major cities, Missoula (Missoula), Gallatin (Bozeman), Yellowstone (Billings), Cascade (Great Falls) and Lewis & Clark (Helena, state capital). The Omicron outbreak in late December begins with large counts in Gallatin (archetype 5) and switches to large counts in all other counties (archetype 6). It ends with significant counts in Cascade and Flathead, but low elsewhere, hence the rise of archetype 3. The sequence of three archetypes in the Omicron outbreak reflects various anti-covid measures put in place during that time period.

**Fig 15.**
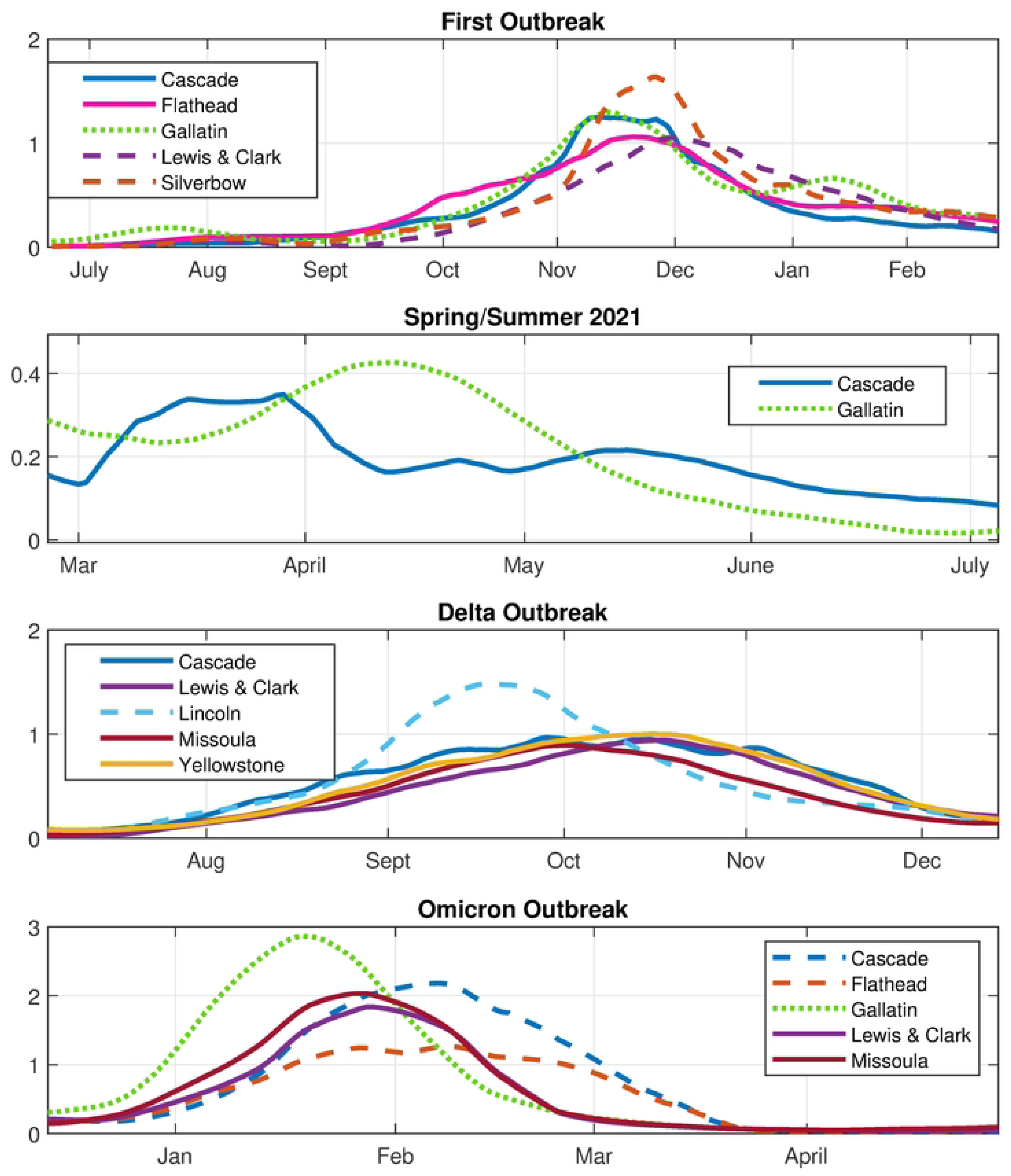
Time series for major component counties in the archetypes featured in different outbreaks. 10 county set.

## Discussion

We have seen how archetypal analysis can be used to good effect in studying a spatio-temporal data set of COVID-19 counts in Montana. Decomposing the entire data set was problematic, however, because of the large size of the normalized counts in small population counties. These small population counties (down to less than 500) have very stochastic signals, and normalized they dominate in the creation of archetypes, which is sensitive to outliers [7]. Hence, we sought ways to remove the stochastic signals of the small population counties. A straight-forward truncation to large population counties with significant city centers formed one reduced set, but we also wanted a way to include the small counties that had significant interaction with the others. To do this, we used a Mutual Information (MI) measure between time series of different counties. A high (relative) MI indicates that a county’s time series can be better predicted by considering the other, and vice versa. We computed the MI between each county and all the others, adding all together for each county to create a total MI, which can be visualized as a spectrum. From this we chose those with the highest total MI, and included all the large population counties, for analysis.

For each presentation of the data, the first archetype is necessarily the zero archetype, which is used to turn off the outbreak in the decomposition of the data. In the 17 county high total MI set we found that certain archetypes were tied uniquely to a given outbreak, while one archetype was used to represent low-level counts in between outbreaks (archetype 2). The initial outbreak was captured by archetypes 5 and 7, which showed a spread from the north central border with Canada and the nascent outbreak in Powell county, to the spread from Powell to the surrounding western counties and from Hill to the eastern counties. Archetype 6 was represents the summer 2020 outbreak which occurred largely in Gallatin county, along with archetype 3, indicating the spread to Cascade county. The Delta outbreak is initialized in archetype 4 which shows counts both in the northwest (Lincoln county), and the far east of the state (Custer county), and ends with archetype 2, which has low-level counts in most counties. The Omicron outbreak was the most complex, as the counts grew and declined several times in different parts of the state, most likely reflecting the reduction in mitigation strategies combined with social gatherings, such as year-end holiday events, and the reconvening of schools in January. COVID-19 first appears in Gallatin county (with its internationally known and large recreational ski area), represented by archetype 6. It then spreads north (Lewis and Clark, Cascade, Hill), east (Yellowstone) and northwest to Missoula, Powell, Lake, Lincoln and Flathead counties, all of which are represented in archetype 8. After that counts decline in many of these counties, lingering more in some counties than others, with a notable late outbreak in Cascade county, hence the later appearance of archetype 9. Finally, it ends with further decline in all counties, with Cascade and Flathead behind the rest, seen in archetype 3.

The 17 county data set shows the influence of small population counties on the eastern and northern border of the state on the initiation and spread of the virus. However, because of of the stochastic signal within small counties and their contribution to outliers in the AA results, we compare this to the Archetypal decomposition of the 10 large population counties data set. It shows the same progression of the epidemic as in the 17 county set, restricted to the high population counties. Archetype 4 for the 10 county data set is similar to archetype 5 in the high total MI set without the low population counties, and both are the main component of the first outbreak. In the Delta outbreak Archetype 2 in the 10 county set is similar to archetype 4. In the Omicron outbreak, Archetype 5 is similar to 8 in the high total MI set, archetype 6 is similar to 9, and archetype 3 to 3. This nesting structure is to be expected, and confirms the validity of the results.

In the 17 county set we see that Archetypal Analysis finds archetypes of counties involved in large outbreaks first, as not including them contributes a sizable amount to the RSS. Therefore, an outbreak may follow a pattern of a low-level outbreak archetype before any hot spots occur, to an archetype with large counts in some selection of counties, perhaps followed by another archetype with another configuration, ending with another low level archetype. The clusters of large outbreak counties is revealed automatically in the analysis, for instance in the initial outbreak a hot-spot in Hill county and larger counts in Powell county spread to all eastern counties and the western side of the state.

How an Archetypal decomposition can be used predicting the spatial spread of disease over time could be explored further. For instance, the information from the Archetypal decomposition could be compared with future outbreaks. Do the outbreaks follow similar spatial patterns? If an outbreak begins in a county that features largely in one archetype, would this imply spread to other counties that have high counts in that archetype? Flathead, Lewis and Clark and Cascade counties have similar outbreak levels in several of the archetypes, which is not surprising, as they are linked by major state roads and have larger population cities, but the analysis confirms that these connections are important. In the east, further analysis could be done focusing on all of the counties in this region to determine how COVID-19 spreads west if initiated in the east. The alpha time series would be used in this analysis, as it represents the transitions from one spatial pattern to another, and holds all time dependent information of the data.

We close by commenting on the care that must be used in the creation and analysis of the archetypes. They have the advantage of automatically showing the counties that experience simultaneous outbreaks, and the state of the other counties during these time periods. Understanding the archetypes is intuitive, unlike graphical representations of PCA vectors. However, if outliers are present in the data (like the inflated counts from small population counties), they can bias the selection of the archetypes. This can be mitigated by filtering out these data points, as we did using an information measure between time series. In doing so we retained the small population counties that were important in the initiation of the outbreaks, as they lie largely on the northern and eastern sides of the state.

## Data Availability

Files will be made available only after acceptance of the ms.

## Acknowledgements

This research was supported by the National Institute of General Medical Sciences of the National Institutes of Health (NIH), United States [Award Number P20GM130418]. We thank the anonymous reviewers for offering feedback on manuscript. We also thank the Montana Department of Public Health and Human Services, Communicable Disease Epidemiology Section, for allowing us access to Montana’s COVID-19 data.

